# Mapping the multidimensional geometric landscape of graded phenotypic variation and progression in neurodegenerative syndromes

**DOI:** 10.1101/2023.10.11.23296861

**Authors:** Siddharth Ramanan, Danyal Akarca, Shalom K. Henderson, Matthew A. Rouse, Kieren Allinson, Karalyn Patterson, James B. Rowe, Matthew A. Lambon Ralph

**Affiliations:** Medical Research Council Cognition and Brain Sciences Unit, University of Cambridge, Cambridge, UK; Department of Clinical Neurosciences and Cambridge University Hospitals NHS Trust, University of Cambridge, Cambridge, UK; Department of Pathology, Cambridge University Hospitals NHS Trust, Cambridge, UK

**Keywords:** heterogeneity, precision medicine, frontotemporal dementia, primary progressive aphasia, machine learning

## Abstract

Clinical variants of Alzheimer’s disease and frontotemporal lobar degeneration display a spectrum of cognitive-behavioural changes varying between individuals and over time. Understanding the landscape of these graded individual-/group-level longitudinal variations is critical for precise phenotyping; however, this remains challenging to model. Addressing this challenge, we leverage the National Alzheimer’s Coordinating Center database to derive a unified geometric framework of graded longitudinal phenotypic variation in Alzheimer’s disease and frontotemporal lobar degeneration. We included three time-point, cognitive-behavioural and clinical data from 390 typical, atypical and intermediate Alzheimer’s disease and frontotemporal lobar degeneration variants (114 typical Alzheimer’s disease; 107 behavioural variant frontotemporal dementia; 42 motor variants of frontotemporal lobar degeneration; and 103 primary progressive aphasia patients). On this data, we applied advanced data-science approaches to derive low-dimensional geometric spaces capturing core features underpinning clinical progression of Alzheimer’s disease and frontotemporal lobar degeneration syndromes. To do so, we first used principal component analysis to derive six axes of graded longitudinal phenotypic variation capturing patient-specific movement along and across these axes. Then, we distilled these axes into a visualisable 2D manifold of longitudinal phenotypic variation using Uniform Manifold Approximation and Projection. Both geometries together enabled the assimilation and inter-relation of paradigmatic and mixed cases, capturing dynamic individual trajectories, and linking syndromic variability to neuropathology and key clinical end-points such as survival. Through these low-dimensional geometries, we show that (i) specific syndromes (Alzheimer’s disease and primary progressive aphasia) converge over time into a de-differentiated pooled phenotype, while others (frontotemporal dementia variants) diverge to look different from this generic phenotype; (ii) phenotypic diversification is predicted by simultaneous progression along multiple axes, varying in a graded manner between individuals and syndromes; and (iii) movement along specific principal axes predicts survival at 36 months in a syndrome-specific manner and in individual pathological groupings. The resultant mapping of dynamics underlying cognitive-behavioural evolution potentially holds paradigm-changing implications to predicting phenotypic diversification and phenotype-neurobiological mapping in Alzheimer’s disease and frontotemporal lobar degeneration.

## Introduction

The challenges of correctly diagnosing, managing and treating Alzheimer’s disease (AD) and frontotemporal lobar degeneration (FTLD) syndromes are exacerbated by phenotypic heterogeneity. Clinically, individuals with AD or FTLD present with diverse cognitive-behavioural profiles and variable disease progression patterns, posing a major challenge to clinical trials and to understanding the mechanisms of syndromic variability. Limited understanding of the landscape of clinical heterogeneity affects diagnostic, stratification and treatment design efforts.^1^ Tackling this challenge first requires mapping the full landscape of graded clinical variations and their dynamic evolution. That is, what are the range and nature of phenotypes present within/between AD and FTLD groups, and how do they change over time? In this work, we take the important step of reconceptualising clinical heterogeneity using a new transdiagnostic multidimensional framework, a large multi-centre dataset, and advanced analytic approaches to map out the first detailed longitudinal picture of graded phenotypic variations in AD/FTLD.

Contextualising this challenge, AD/FTLD are not monomorphic entities; instead, they comprise clinical variants that, at least in early stages, differentially affect episodic memory (typical amnesic AD),^2^ language (primary progressive aphasia, PPA),^3^ motor function (Corticobasal Syndrome, CBS; Progressive Supranuclear Palsy, PSP),^4,5^ semantic cognition (semantic dementia/semantic variant PPA),^6,7^ behaviour and/or executive functions (behavioural variant frontotemporal dementia, bvFTD).^8^ Typically, variants are diagnosed as independent entities based on discrete symptoms and corresponding neural-biological dysfunction profiles. In the reality of everyday clinics, there are many exceptions to such ‘textbook’ cases. Patients frequently present with, and evolve to show, mixed symptom profiles spanning multiple independent diagnostic categories, or display spared/fewer features leading them to miss key diagnostic criteria.^9^ This phenotypic variation can emerge independent of disease severity,^10,11^ is systematically present across variants,^9,12–14^ and relates to underlying brain structure-function integrity^15,16^ and genetic-pathological mechanisms.^17–19^ Mapping these variations is of significant clinical and research importance as: (i) initial clinical diagnosis is predicated on cognitive-behavioural profiles; (ii) heterogeneous individuals challenge current nosology, risking exclusion from clinical trials;^20^ (iii) many dementia screening programmes probe for prototypical clinical profiles, leading to missed diagnosis/misdiagnosis of heterogeneous cases, in turn questioning their accuracy;^21^ (iv) insufficient understanding of phenotypic heterogeneity precludes confident prediction of disease trajectories/endpoints and tailored management and care information provision;^22^ and (v) excluding heterogeneous cases from research studies represents a major missed opportunity to reveal systematic transdiagnostic variations or universal symptoms and their neurobiological bases, and to understand moderators of disease presentation or progression.^23^ Considering clinical variations in terms of categorical distinctions (and inherent sources of data noise) is a common approach. Such frameworks are challenged, however, when there is considerable variation within as well as between categories, in the limit undermining the very presence of the categories themselves. Rather than solely conceptualising clinical variations as emerging from disease-linked categorical generators, modelling along continuous dimensions offers an additional robust explanatory framework of heterogeneity.^24,25^ Dimensional variations are commonly found both in the natural world (e.g., temperature, light, pressure) and in general medicine (e.g., blood pressure, glycaemic control, renal function, body mass index). In AD/FTLD, a colour analogy offers an intuitive explanation of modelling heterogeneity using dimensional frameworks.^16^ Consider two classic exemplar clinical conditions (e.g., semantic dementia vs. typical amnestic AD); these can be thought of as strongly contrastive, or even “complementary” colours (e.g., yellow vs. blue) within a continuous, multidimensional hue-space. Systematic shifts along one or more dimensions generate not only variations around these exemplar cases (e.g., lemon, mustard, Chartreuse), but also “mixed” cases reflecting combinations of the core dimensions (e.g., orange, lime). Dimensional frameworks have the potential to represent longitudinal changes in terms of movements along one or more disease-linked dimensions, that otherwise confusingly appear to be multiple changes in diagnosis (category; e.g., orange→coral→salmon→pink→fuchsia simply reflects a blue dimensional shift). Expanding this example to the wider range of AD/FTLD phenotypes – instead of defining an ever finer-grained list of category subtypes (cf. the mesmerising array of names on a paint colour chart), dimensional approaches distil performance covariance patterns to unveil the fundamental phenotypic dimensions (cf. colour axes) on which patients differ, mapping graded clinical variations and multifaceted longitudinal changes (cf. fuzzy boundaries between hues/shades) to a unified multidimensional phenotypic geometry (cf. hue space). Of note, considering a dimensional approach does not mean discounting or abandoning categorical approaches to clinical labelling. Categorical labels do hold clinical utility and, while undeniably imperfect like any other approach, they are helpful for explaining the nature and the course of the disease to patients. Dimensional approaches offer a complementary fine-grained underpinning framework that enables precision medicine, with precision not in terms of gene or molecule but in terms of the actual clinical syndrome.

This perspective offers several potential advantages in linking transdiagnostic clinical variability to common and diverse neurocognitive, brain, molecular and pathological axes of changes, all within one unified framework. Accordingly, these approaches are increasingly popular within psychiatry,^25^ neurodevelopment,^26,27^ post-stroke aphasia^28,29^ and recently adopted in neurodegeneration research.^10,16,30–32^ When applied to AD/FTLD, we can better (i) understand key latent mechanistic drivers of structured phenotypic heterogeneity; (ii) co-locate and inter-relate typical, intermediate and atypical clinical presentations; and (iii) visualise individual disease trajectories, revealing longitudinal phenotypic convergence or divergence of syndromes. Furthermore, these phenotype geometries readily relate to everyday clinic observations to explain variation across canonical presentations and clinical “chameleons”^33^ such as bvFTD with AD-like severe amnesia^34,35^ or FTLD-motor syndromes with aphasia,^36^ accommodate locations of broad categorical descriptors such as “mixed aphasia” or “atypical dementia”, and relate them to different neural-pathological mechanisms.^31,32,37,38^ In short, leveraging phenotypic heterogeneity to clinical advantage remedies limitations of categorical-only descriptive approaches, models fluid intersections and dynamic trajectories of syndromes, and reveals the large landscape of nuanced inter-individual clinical variations in AD/FTLD. In turn, this approach may further help: to yield reliable markers identifying particular disease processes amid the wider phenotypic spectrum; to specify certain features (e.g., behavioural change) that are understated/emerge with time that may be otherwise overlooked; to aid prognosis of individual cases; and ultimately to guide the treatment of patients.

In this work, we apply such a data-driven approach to large-scale longitudinal AD/FTLD clinical and cognitive-behavioural data to unveil a multidimensional geometry of phenotypic variation. We constructed these geometries in a group of sporadic AD and FTLD typical, atypical and intermediate clinical variants (*N*=390) from the large National Alzheimer’s Coordinating Center (NACC) database. All patients were assessed at three consecutive time points on 39 clinical measures spanning behaviour, cognition, motor ability, personality, mood, and psychiatric changes. Given the range of measures and broad sampling of the AD-FTLD syndromes, the resultant space maps the large landscape of early-to-moderate clinical symptoms, their varying severity, and transdiagnostic frequency. On these data, we applied new analytic approaches adopted from data-science, machine learning, contemporary applied mathematics, chemometrics, and geostatistics. Specifically, we first used varimax-rotated principal component analysis (PCA), a long-established method to derive multiple intuitive orthogonal phenotypic axes linked to recognisable clinical variations, thereby capturing patient-specific movement along and across these dimensions. Then, we adopted more recent non-linear neighbour embedding methods (Uniform Manifold Approximation and Projection; UMAP) to distil the PCA axes and patient locations into a visualisable 2D manifold of longitudinal phenotypic variation. This step helped to map and visualise parallel and non-linear patterns of progression simultaneously along the multiple PCA dimensions, at the level of the individual and group. By projecting individual points into this geometry, we derived key insights such as homophily/heterophily with disease progression (i.e., within-group similarity/differentiation), convergence of syndromes into a “de-differentiated pooled phenotype”, clinical variations in those with unstable diagnoses, and associations with clinico-pathological endpoints such as survival and pathological status. This final step revealed phenotypic predictors of tissue pathology, with potential to inform treatment decision-making for patients based on clinical presentation. This unified visualisable framework of graded syndromic/symptomatic variations offers an unprecedented view into the dynamics of cognitive-behavioural evolution in AD/FTLD, concurrently at individual- and group-levels.

## Methods

### Participants

We derived the sample from the December 2021 data freeze of the Uniform Data Set (UDS; for visits conducted between June 2005-November 2021) of the NACC dataset (https://naccdata.org/).^39^ Our analyses used data from 17 Alzheimer’s Disease Research Centers (ADRCs). We retained patients with either typical, intermediate, or atypical clinical diagnoses of AD or FTLD (all sporadic cases) whose clinical diagnosis was rated as the primary cause of their cognitive impairment (i.e., dementia). Participants were excluded if they had a clinical diagnosis of Dementia with Lewy Bodies, Motor Neurone Disease, Mild Cognitive Impairment or subjective cognitive complaints/worried well, any other primary or secondary neurological disorder contributing to their cognitive status, Down’s syndrome, substance abuse, primary psychiatric illnesses (e.g., depression, schizophrenia, bipolar disorder), delirium, traumatic brain injury, normal pressure hydrocephalus, vascular brain impairment, and/or multiple sclerosis.

The final sample included 390 patients, all diagnosed as per current clinical diagnostic criteria: 114 AD,^2^ 107 bvFTD,^8^ 8 PSP^5^ and 34 CBS^4^ (grouped together and referred to as FTLD-motor), 24 FTLD not otherwise specified (NOS, comprising mixed clinical presentations of FTLD syndromes), and 103 PPA^3^ (7 semantic variant PPA, 18 logopenic variant PPA, 13 nonfluent variant PPA, 9 PPA-NOS and 57 “PPA”, with the latter two comprising mixed clinical presentations of PPA). For PPA, specifically, we note that the disproportionately greater number of mixed cases without subtype designation, as compared to canonical variants, is a characteristic of the database also reported by others^40^ (see Supplementary Methods, Supplementary Tables 1-3 and Supplementary Figure 1 for a breakdown of the PPA group by recruitment site, year of entry into the data cut, and, for the mixed PPA patients, their corresponding diagnostic label per the older criteria from Mesulam^41^). All clinical diagnostic labels were already provided for each patient in the NACC database. Following the first visit, 88 patients (2 AD, 38 bvFTD, 13 FTLD-motor, 2 FTLD-NOS, and 33 PPA) had their initial clinical diagnosis changed over one of the two consecutive assessments (Supplementary Figure 2), shifting between diagnostic categories (e.g., bvFTD to FTLD-motor) or receiving a more specific diagnosis within a category (e.g., “PPA” to semantic variant PPA). These individuals were deliberately included in the full sample to model evolution between fuzzy syndrome boundaries. All selected patients had three consecutive time-point data (total data points = 1,170) including their first reported NACC entry (baseline/Visit 1) and two consecutive follow-ups (Visits 2 and 3) assessed every 1.2 years on average.

### Behavioural and cognitive measures

We chose 39 performance and rating measures spanning a range of clinical, cognitive, behaviour and mood metrics, carefully selected to represent a broad range of measured functions and following checks for minimal missing data, distribution patterns and floor/ceiling effects. Final behavioural measures of interest that were entered into the main analyses included (i) select subdomains of the Neuropsychiatric Inventory^42^ (Motor, Irritability, Hallucinations, Elation, Disinhibition, Depression, Appetite, Apathy, Anxiety, and Agitation); (ii) select subscores of the Geriatric Depression Scale^43^ (Worthlessness, Hopelessness, Helplessness, and Emptiness in life); (iii) clinician rated scores on changes in motor function (tremors, slowing, gait, falls), behaviour (visual and auditory hallucinations, delusions, depression, disinhibition, irritability, personality changes) and cognitive status (memory, judgment/planning/executive function), (iv) verbal fluency (animals, vegetables); and (v) subdomains of the Functional Activities Questionnaire^44^ (changes in capacity to travel, pay taxes, operate the stove, shop, remember dates, pay attention, prepare meals, play games, attend events, and pay bills). To validate the statistical structure of our UMAP analyses and relate it to clinical observations, we further selected three independent variables that were not used in any part of our the main analytic pipeline - clinician rated disease severity (Clinical Dementia Rating Plus NACC FTLD-Sum of Boxes, CDR-FTLD-SoB)^45,46^ and measures of predominant changes in cognition (*NACCOGF*) and behaviour (*NACCBEHF*) at each visit.

### Pathology classification

From the full patient group (*N*=390), *N*=209 (~53%) had reported deceased status. Of this sample, *N*=100 patients (47%) died within 3 years of their first recorded NACC visit (Supplementary Figure 3). Of the total sample with reported deceased status, *N*=139 (~66%; 24 AD, 49 bvFTD, 26 FTLD-motor, 3 FTLD-NOS, 37 PPA) patients had available pathological information. For these individuals, we extracted their pathological information from the NACC Neuropathology Data Set and respective details from the Neuropathology Data Dictionary. It should be noted that, for many individuals, pathological information was coded and scored differently in previous versions of the NACC Neuropathology Data Set;^47^ for these cases, primary pathological diagnosis is indicated with variables with the prefix *NP* and this information was used as the primary pathological diagnosis. For others, through discussions with an expert neuropathologist (K.A.) and neurologist (J.B.R.), we computed their primary pathological diagnosis using other relevant columns, described below.

- Primary Alzheimer’s disease pathology was coded using the *NPPAD* variable and/or the NIA-AA Alzheimer’s disease neuropathologic change score (*NPADNC* variable) to categorise low, intermediate and high Alzheimer’s disease neuropathologic change.^48^
- Primary cerebrovascular disease (primary vascular pathology) was coded using the *NPPVASC* variable. With inputs from an expert neuropathologist (K.A.) and neurologist (J.B.R.), we stratified co-occurring vascular changes into 4 categories (NACC codes for each of these variables are detailed in Supplementary Table 4):

`o` amyloid angiopathy (cerebral amyloid angiopathy)
`o` non-amyloid angiopathy
`o` vascular changes (large arterial infarcts, one/more lacunes, microinfarcts, arteriosclerosis, subcortical arteriosclerotic leukoencephalopathy, white matter rarefaction, other pathological changes related to ischemic or vascular disease not previously specified, laminar necrosis, mineralization of blood vessels, and/or other ischemic/vascular pathology)
`o` acute injury (single/multiple haemorrhages, cerebral microbleeds, acute neuronal necrosis, and all acute/subacute gross infarcts, microinfarcts, gross haemorrhages and/or microhaemorrhages).
`o` We excluded information on any type of aneurysm, vasculitis, vascular malformations, and cerebral autosomal dominant arteriopathy with subcortical infarcts and leukoencephalopathy (CADASIL).
- Lewy Body Disease pathology was coded using the *NACCLEWY* and *NPLBOD* variables.
- Progressive Supranuclear Palsy pathology was coded using the *NACCPROG* variable.
- Corticobasal Degeneration pathology was coded using the *NACCCBD* variable.
- FTLD-Pick pathology was coded using the *NACCPICK* variable.
- FTLD-Ubiquitin pathology was coded using the *NPFTD* variable.
- FTLD-Tau pathology was assigned based on “present” value for FTLD with tau pathology or other tauopathy (*NPFTDTAU*) or FTLD-tau subtypes including other 3R/4R and 3R+4R tauopathies (*NPFTDT2*, *NPFTDT6*, *NPFTDT10*), argyrophilic grains (*NPFTDT5*), tangle dominant disease (*NPFTDT9*), frontotemporal dementia and parkinsonism with tau-positive or argyrophilic inclusions (*NPFRONT*), and other tauopathy (*NPTAU*). We excluded FTLD-tau subtype chronic traumatic encephalopathy and amyotrophic lateral sclerosis/parkinsonism-dementia pathologies.
- FTLD-TDP-43 pathology was assigned based on “present” value for FTLD with TDP-43 pathology (*NPFTDTDP*) including TDP type A/B/C/D/E (*NPTDPA/B/C/D/E*).
- FTLD-Other pathology was coded using the *NPOFTD* variable (other FTLD) and/or the *NPOFTD5* (FTLD-NOS includes dementia lacking distinctive histology and FTLD with no inclusions detected by tau, TDP-43, or ubiquitin/ph62 immunohistochemistry). We excluded atypical FTLD-U, neuronal intermediate filament inclusions disease, basophilic inclusion body disease, and FTLD-ubiquitin-proteasome system pathologies.
- Other pathology was coded based on “present” value for pigment-spheroid degeneration/NBIA (*NPPDXA*), white matter disease – leukodystrophy (*NPPDXG*), and metastatic neoplasm (*NPPDXL*) and other pathologic diagnosis (*NACCOTHP*).

We excluded primary pathologic labels of amyotrophic lateral sclerosis/motor neurone disease, prion disease, multiple system atrophy, cortical development malformation, metabolic/storage disorders, demyelinating white matter diseases (multiple sclerosis), traumatic brain injury/contusion, and infectious diseases.

We also note that a significant percentage of individuals with available pathological data had co-occurring vascular changes. In two cases, there was clear indication of primary cerebrovascular pathology whereas for the others, information on the primacy of vascular contribution was unavailable. For brevity, we therefore (i) report the frequency and co-occurrence of these vascular changes but do not focus on their magnitude or contributions, and (ii) do not focus on the frequency and contributions of other co-pathologies as it is beyond the scope and interest of this study.

### Statistical analyses

Statistical analyses were conducted in RStudio v.4.2.0^49^ and MATLAB.^50^ A full list of R packages, their primary uses, and associated references are listed in Supplementary Methods.

#### Group differences on clinical and demographic performance

For binomially-distributed variables (sex), chi-squared tests were used. For continuous variables (age, education, disease severity, duration between follow-ups), we conducted analyses of variance (ANOVA) with post-hoc Tukey’s Honest Significant Differences corrections for multiple comparisons, accompanied by effect sizes (eta-squared, η^2^) and 95% confidence intervals.

#### Principal component analysis

Behavioural and cognitive measures were ordered in the same direction, scaled to percentages, and missing data (5.9% overall) were imputed using *mice* regression models with age, education, sex, diagnosis, visit number and disease severity (CDR-FTLD-SoB) as predictors, per previous protocols.^51^ A key interest of this study was to map covariance structures over time; therefore, we determined the number of PCs underlying this dataset, both irrespective of and accounting for visit number. Scree plots, parallel analysis and 4-fold cross-validated PCA component selection (an approach used in chemometrics)^52^ were used to estimate the number of PCs underlying the dataset. It is noteworthy that all methods converged to suggest six PCs irrespective of time (Supplementary Figure 4), with the same factor structure emerging irrespective of/even when data were stratified by time.

The full dataset (*N*=1,170) was entered into an orthogonally-rotated (varimax) PCA, which extracted six components, each representing a separate source of variation underlying the data. Orthogonal rotations allow for minimal inter-component shared variance by maximising loading dispersion, therefore promoting clear behavioural interpretability. Measures loading >|.5| were treated as important contributors and components were labelled for ease of reference; however, we note labels function as short-hands and may not fully capture all measures loading on a given component. PC scores across all six PCs were considered together to represent a “multidimensional PC space” where each patient’s location is a proxy for their overall cognitive-behavioural profile at that time point.

#### Distance-based analysis within multidimensional PCA space

The multidimensional PC space afforded the unique capacity to probe the direction, magnitude, and nature of symptom changes. These analyses were conducted in three steps.

First, we probed the direction of movement to ask “how phenotypically similar do neurodegenerative disorders become over time?” We calculated a global dispersion index to quantify how much a person looks like/unlike other neurodegenerative dementia syndromes over time. Using this metric, we specifically tested if patients (i) irrespective of diagnosis, became increasingly similar to each other over time to converge globally (“de-differentiation” from pooled phenotype hypothesis), (ii) became more similar within-group but dissimilar between-groups (“differentiation” from pooled phenotype hypothesis); and/or (iii) displayed negligible changes between each other from baseline (“stable from baseline” hypothesis). For each data point, we calculated its distance from the centroid of the multidimensional PC space to form a global dispersion index. Over time, movement towards the centroid supports the “de-differentiation hypothesis” where syndromes converge towards a global amalgamate and eventual clustering of individuals around a de-differentiated pooled/generic dementia phenotype, whereas divergence from the centroid indicates differentiation from the group aggregate and syndromes remaining true-to-type with disease progression. Changes on the global dispersion index were assessed using linear mixed-effect models with a fixed effect for group and follow-up time, a random effect for follow-up time, a random intercept for individual performance, and a random slope for follow-up time (hereon, referred to as the standard linear mixed-effect model pipeline for brevity). We repeated the global dispersion index analyses in individuals with stable vs. changing diagnostic labels with the same pipeline. For the analysis in those with changing diagnostic labels, we added an additional random effect of group. Finally, two-tailed Pearson’s correlations were used to examine strength of associations between global dispersion index and disease severity (CDR-FTLD-SoB).

To quantify the magnitude of phenotypic change in each patient group, we asked “which groups show the greatest cognitive-behavioural changes over time?” To do so, we calculated the Euclidean distances between each individual’s baseline and follow-up data points and examined within-group differences using ANOVAs with planned Tukey’s comparisons.

To explore the nature of phenotypic change in each patient group, we repeated the dispersion index calculation within each individual PC to derive a PC-specific dispersion index (i.e., distance from each PC’s centroid). On this value, we refitted our standard linear mixed-effect model pipeline. These PC-specific dispersion were used again in analyses of associations with survival patterns.

#### Manifold learning with UMAP

The multidimensional PC space had six independent linear phenotypic axes underpinning the cognitive-behavioural heterogeneity and longitudinal progression of AD/FTLD. However, this 6D space is challenging to visualise and to infer parallel and non-linear progression patterns simultaneously across multiple dimensions. To address this, we used manifold machine-learning, specifically UMAP,^53^ to visualise and map longitudinal interdigitation and diversification of AD/FTLD. UMAP learnt the manifold structure of the 6D PC space, reconstructed a high-fidelity 2D embedding,^54^ preserving the topological structure of the original high-dimensional space (through non-linear dimension reduction on a Riemannian manifold)^55^ whilst clustering or dispersing data points based on their similarities (irrespective of time).^56,57^ In learning the topographic structure of the data, UMAP can drive groups and individuals apart if they are fundamentally dissimilar, proving a way to map phenotypic similarity concurrently at the level of the individual and group (within a “UMAP space”). The resultant curved manifold can be projected onto a flat plane, much like mapping a globe onto a flat map. Like geographical maps, this means that in central parts of the 2D plot, points which are closer to each other tend to be more similar (e.g., Portugal is closer to Spain than Greece). However, outer parts of the projection may appear far away from each other, but in fact may be actually closer (e.g., Canada and Russia). Together, in the “UMAP space”, inter-relation of distances and positions of data points provide a low-dimensional interpretable structure to longitudinal movement of patients.^57–60^

Per standard recommendations, we initialised the UMAP using the PCA data.^61,62^ We tuned two key hyperparameters regulating the distance between points, position of points, and clusters in the data: number of neighbours (here, the default 15), and minimum distance (here, the default 0.1). Briefly, the ‘number of neighbours’ hyperparameter constrains the number of neighbouring points considered when analysing the data in low-dimensional space. Lower ‘number of neighbours’ values force the UMAP to concentrate on a highly local structure while larger values push points away from each other to focus on a global neighbourhood at the cost of giving up finer details, analogous to the community structure of a village street vs. a large residential community. The minimum distance parameter controls the minimum distance between the points in the low-dimensional space. Smaller values result in clumpier embeddings of smaller connected components, whereas larger values give an overarching view of the data at the cost of detailed topological structure.^53^ Both parameters together give a balanced view of global vs. local structure of the data. Returning to our colour analogy, larger ‘number of neighbours’ and ‘minimum distance’ values would capture the overall structure of colours (i.e., blues, greens, yellows, high vs. low luminance) but at the cost of finer local structure where individual colours may not be necessarily close to their nearest colour match. The hyperparameter values chosen in the current study provide a good balance of local and global structure relevant to the dataset and have been previously used in data of similar structure.^58,63^ Within the UMAP space, we mapped spatial locations and density of each group using 2D kernel density estimation (KDE) plots. We tested statistical differences between densities stratified by time and group using spatiotemporal 2D KDE statistics.

Finally, we visualised animated movements of each individual point within the UMAP space (see https://github.com/siddharthramanan/NACC_UMAP/tree/main/UMAP_animate).

#### Testing the statistical structure of data distribution within the UMAP space

It is important to understand whether the UMAP was able to understand the spread of data and pick out overlaps and differences. We tested the structure of the distribution to uncover whether the UMAP embedding was completely random or reflected meaningful graded co-location of individual data points and patient groups. We borrowed methods from geospatial statistics, namely spatial point pattern analysis, average nearest neighbours (ANN, computed in Euclidean distances) and Monte Carlo simulations. We then created a “null model” of 1,170 randomly distributed points (equivalent to the number of data points embedded within our UMAP space) embedded within the convex hull of our UMAP distribution. We then ran 1,000 simulations to extract mean ANN differences between null and observed data, computed *p*-values, and plotted distribution histograms.

Respecting the location of individual data points and patterning within the UMAP space, we tested whether patient movements within this space was indicative of phenotypic convergence or dispersion to de-differentiate. Within groups, we stratified the data by time, and repeated our previous ANN pipeline with one critical difference in null model creation. Instead of simulating a truly random distribution as the null model, we created null models by randomly shuffling group labels within each time-stratified dataset (creating a null model equivalent to the *N* of each stratum). Such a null model respected patterning within the UMAP space (as tested in the aforementioned analyses) but simulated a null distribution where patient groups may locate at different points within the UMAP that are currently occupied by a member of a different clinical group. All null models were sampled equal to the observed distribution. We then repeated the ANN analyses (1,000 simulations), compared the ANN values from null and observed data, and plotted these histograms.

#### Relating manifold spaces to clinical profiles

The under-the-hood transformations of UMAP can create a ‘black box’ situation, where there is limited direct correspondence between each individual’s raw data and its expression as a coordinate in the UMAP space.^56^ To facilitate this interpretability, we derived clinically-meaningful information from the UMAP space by linking it back to our multidimensional PC space and other clinical data. We rank ordered each PC score from highest (best performance) to lowest (worst performance), divided these into tertiles, and projected both ranks (as size-varying dots with larger points=lower rank/worse performance) and tertiles (as 2D KDE contours) into the UMAP space. We also projected *NACCCOGF* and *NACCBEHF* variables as dot plots into the UMAP space to understand the location of patients reporting specific cognitive and behavioural complaints, respectively. As these variables were not included in the computation of the PCA/UMAP, they also serve as independent validators of symptom-syndrome-location mapping within the embedding. We further created an interactive plot of key demographic and disease-related values for each individual at their given UMAP location over time <u>(see attached Supplementary HTML UMAP Plotly file</u>).

#### Predicting survival status from location and movement in multidimensional phenotypic spaces

Whether transdiagnostic or variant-specific features powerfully predict pathological status and key clinical endpoints such as survival remains less well understood. As ~53% (*N*=209) of the sample had reported deceased status, this study was uniquely placed to explore the relationships between clinical heterogeneity, survival and pathological status in AD/FTLD.

First, we sought to understand whether the multidimensional PC and UMAP spaces could inform of key clinical end-points, namely survival at 36 months. In 209 patients who had reported deceased status, we classified these individuals as “deceased” if they had reported deceased status within the 36 months of their first recorded NACC visit (*N*=100), else “not deceased”. We then examined the associations with symptom severity between individuals who were deceased within 36 months, deceased after 36 months, and those who were not deceased. Next, focusing on global metrics (global dispersion index in the multidimensional PC space) and local metrics (PC-specific dispersion indices), we revisited our global and PC-specific dispersion indices and, for each metric, used the baseline values as “starting points” (indicating how impaired someone is to start with) and average dispersion values over all time points as “travelled distance” (indicating how much this impairment evolves with time). Distinguishing starting points and travelled distance respectively models effects of having a head-start in global cognitive-behavioural performance and subsequently variable patterns of decline, independent of baseline performance.

For the multidimensional PC space, we used starting points and travelled distances from the global dispersion index and entered these into multivariate stepwise logistic regressions, with models with best fit evaluated using the Akaike Information Criterion (AIC) parameter.

For the UMAP space, we restricted the model to baseline data, treated each person’s UMAP location as a spatial coordinate, and used generalised linear models, and non-linear spatial regression models from geostatistics to predict survival at 36 months. For the linear model, we modelled the scaled interaction of *x* and *y* UMAP coordinates on survival at 36 months. For the non-linear model, we used generalised additive models with a smoothed interaction of *x* and *y* UMAP coordinates predicting survival at 36 months. The generalised additive model included a thin plate spline, with smoothing done through Gaussian process model optimisation, and Restricted Maximum Likelihood parameters.

For PC-specific dispersion indices, we built group-wise multivariate stepwise logistic regression models entering starting points and travelling distance values for all PCs as predictors of survival status at 36 months.

Models with best fit were evaluated based on the AIC parameter, which inherently adds a penalty term for the complexity of the model.

#### Associations between low dimensional spaces and pathological grouping

Turning to pathology data, 139/209 (~66%) reported deceased patients had histopathological data that we grouped under 11 broad pathological brackets: primary AD pathology, seven different FTLD pathologies, cerebrovascular disease (CVD), Lewy Body Disease (LBD), and Other pathologies. Most patients with clinical AD, bvFTD, FTLD-motor and PPA diagnoses had co-occurring vascular changes (Supplementary Figures 5-6). For brevity and simplified interpretation, we focus on the 11 major pathological groupings and do not consider co-occurring vascular changes in the following analyses (except for 1 bvFTD and 1 PPA where CVD was the primary pathological diagnosis).

We then projected primary pathology data into the UMAP space and tested whether, irrespective of time, pathological groups are situated in clusters or spread randomly. To test whether spatial embedding of pathological groups was random or patterned, we repeated the aforementioned spatial point pattern and ANN analysis by creating null models sampled equivalent to the *N* of each pathological group embedded within the convex hull of the UMAP space, running 1,000 simulations to quantify differences between null and observed data for each pathological group, computing *p*-values, and plotting distribution histograms.

To examine longitudinal changes in PC scores between the 11 pathological grouping labels, we conducted simple linear regressions modelling the interaction of group and time on each PC score. We did not employ linear mixed-effect models as, for the standard linear mixed-effect model pipeline, the number of observations was significantly lesser than the number of random effects making the random-effects parameters and the residual variance (or scale parameter) unidentifiable.

### Data and code availability

The National Alzheimer’s Coordinating Center dataset are freely available through request on their official website (https://naccdata.org/). Code for all analyses from this study have been made available at: https://github.com/siddharthramanan/NACC_UMAP.

## Results

### Sample characteristics

Baseline demographic and clinical variables information are displayed in Table 1. At baseline, group comparisons on demographic variables revealed a significant effect of sex (χ =23.5; *p*<.001) with more females in AD group, and more males in the bvFTD and FTLD-NOS groups (all *p*<.001). AD patients were significantly older and had fewer years of education compared to other groups (all *p*<.02). AD and bvFTD had significantly greater clinician-indexed disease severity (CDR-FTLD-SoB) compared to PPA (both *p*<.01). Additionally, the bvFTD group had significantly greater disease severity compared to FTLD-motor patients (*p*<.01). These patterns mirror previously reported population statistics.^64,65^ No significant group differences were noted for symptom duration (*p*>.1). Annual follow-ups occurred approximately every 1.2-1.4 years, with no statistical differences between groups on inter-assessment interval (all *p*>.08).

**Table 1.**
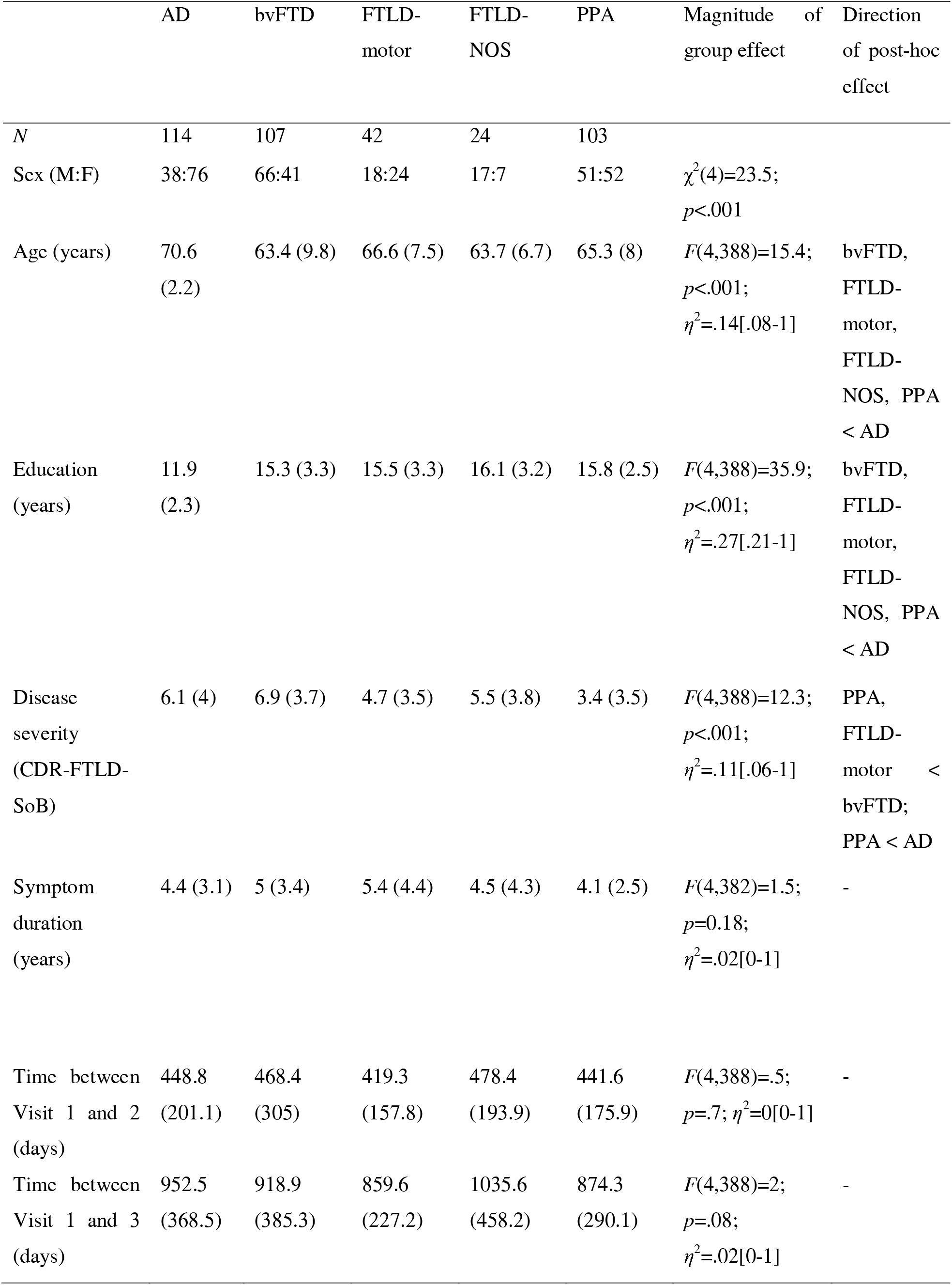
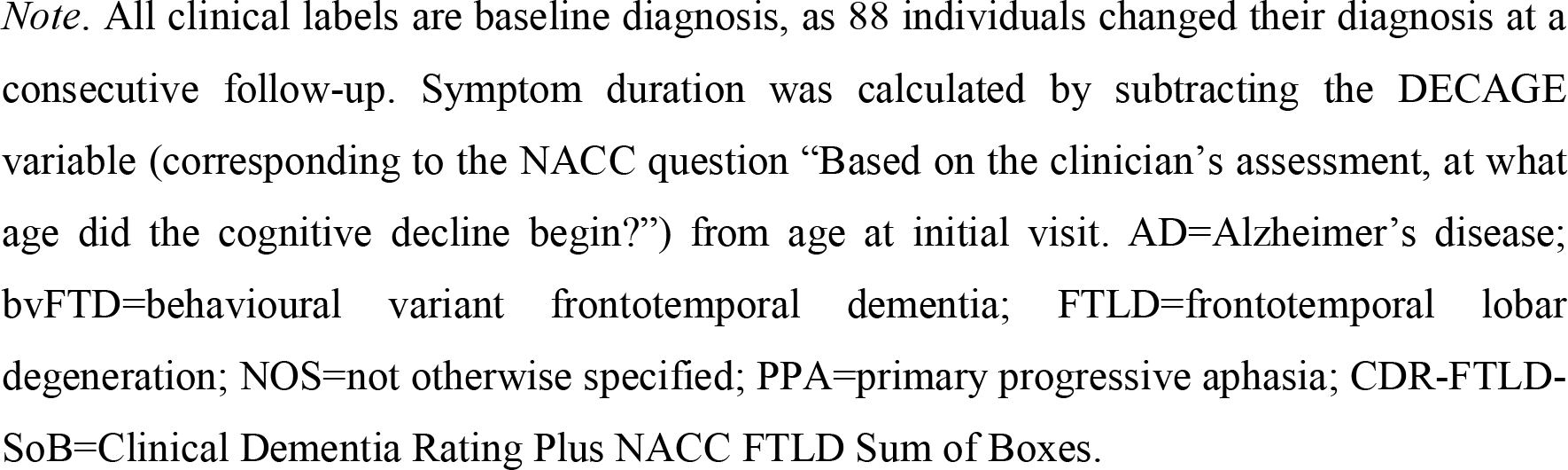
Baseline demographic and clinical characteristics of participant groups.

|  | AD | bvFTD | FTLD-motor | FTLD-NOS | PPA | Magnitude<br>of group effect | Direction<br>of post-hoc<br>effect |
| --- | --- | --- | --- | --- | --- | --- | --- |
| *N* | 114 | 107 | 42 | 24 | 103 |  |  |
| Sex (M:F) | 38:76 | 66:41 | 18:24 | 17:7 | 51:52 | $\chi^2(4)=23.5$;<br>$p<.001$ | |
| Age (years) | 70.6<br>(2.2) | 63.4 (9.8) | 66.6 (7.5) | 63.7 (6.7) | 65.3 (8) | $F(4,388)=15.4$;<br>$p<.001$;<br>$\eta^2=.14[.08-1]$ | bvFTD,<br>FTLD-motor,<br>FTLD-NOS, PPA<br>< AD |
| Education<br>(years) | 11.9<br>(2.3) | 15.3 (3.3) | 15.5 (3.3) | 16.1 (3.2) | 15.8 (2.5) | $F(4,388)=35.9$;<br>$p<.001$;<br>$\eta^2=.27[.21-1]$ | bvFTD,<br>FTLD-motor,<br>FTLD-NOS, PPA<br>< AD |
| Disease<br>severity<br>(CDR-FTLD-SoB) | 6.1 (4) | 6.9 (3.7) | 4.7 (3.5) | 5.5 (3.8) | 3.4 (3.5) | $F(4,388)=12.3$;<br>$p<.001$;<br>$\eta^2=.11[.06-1]$ | PPA,<br>FTLD-motor <<br>bvFTD;<br>PPA < AD |
| Symptom<br>duration<br>(years) | 4.4 (3.1) | 5 (3.4) | 5.4 (4.4) | 4.5 (4.3) | 4.1 (2.5) | $F(4,382)=1.5$;<br>$p=0.18$;<br>$\eta^2=.02[0-1]$ | - |
| Time between<br>Visit 1 and 2<br>(days) | 448.8<br>(201.1) | 468.4<br>(305) | 419.3<br>(157.8) | 478.4<br>(193.9) | 441.6<br>(175.9) | $F(4,388)=.5$;<br>$p=.7$; $\eta^2=0[0-1]$ | - |
| Time between<br>Visit 1 and 3<br>(days) | 952.5<br>(368.5) | 918.9<br>(385.3) | 859.6<br>(227.2) | 1035.6<br>(458.2) | 874.3<br>(290.1) | $F(4,388)=2$;<br>$p=.08$;<br>$\eta^2=.02[0-1]$ | - |

*Note.* All clinical labels are baseline diagnosis, as 88 individuals changed their diagnosis at a consecutive follow-up. Symptom duration was calculated by subtracting the DECAGE variable (corresponding to the NACC question “Based on the clinician’s assessment, at what age did the cognitive decline begin?”) from age at initial visit. AD=Alzheimer’s disease; bvFTD=behavioural variant frontotemporal dementia; FTLD=frontotemporal lobar degeneration; NOS=not otherwise specified; PPA=primary progressive aphasia; CDR-FTLD-SoB=Clinical Dementia Rating Plus NACC FTLD Sum of Boxes.

### AD and FTLD clinical heterogeneity vary along six principal axes

In the full dataset (*N*=1,170), we first explored the landscape of longitudinal phenotypic heterogeneity by constructing a multidimensional geometry of transdiagnostic performance changes on test measures using varimax-rotated PCA. The feature:sample ratio was adequate (Kaiser-Meyer-Olkin statistic=.88; Supplementary Figure 7). Our component selection methods converged on a 6 PC solution (explaining 51% of overall variance; Supplementary Figures 4 and 8) with the same factor structure, irrespective of lumping or stratifying the dataset by time. PCA solutions and explained variance are direct reflections of the homogeneity of the sample and input variables. It is encouraging, therefore, that the PCA resulted in six clinically-intuitive principal components and this solution explained over half of the overall variance, especially considering that the input features comprised heterogeneous assessments and a highly mixed sample of canonical and atypical variants evolving across three distinct time points with considerable individual differentiation.

PC1 (labelled “Functional status”) explained 19% of this variance, loading positively on measures of functional activities, clinician-indexed memory and executive performance, and verbal fluency (Figure 1A). PC2 (“Apathy/impulsivity”) explained 9% of overall variance and loaded positively on Elation, Apathy, Disinhibition, Motor, Appetite subdomains of the Neuropsychiatric Inventory (NPI), and clinician-indexed disinhibition and personality change scores. PC3 (“Motor function”) explained 7% of overall variance and loaded positively on clinician-indexed changes in gait, falls, tremors and slowing of movement. PC4 (“Psychosis”) explained 6% variance and loaded positively on NPI Hallucinations subdomain and clinician-rated scores on auditory/visual hallucinations and delusions. PC5 (“Affective changes”) explained 5% of variance and loaded positively on NPI Agitation, Depression, Anxiety and Irritability subdomains, and clinician-scored Depression and Irritability. PC6 (“Depression”) loaded on Geriatric Depression Scale and explained 5% of overall variance (Figure 1A).

**Figure 1.**
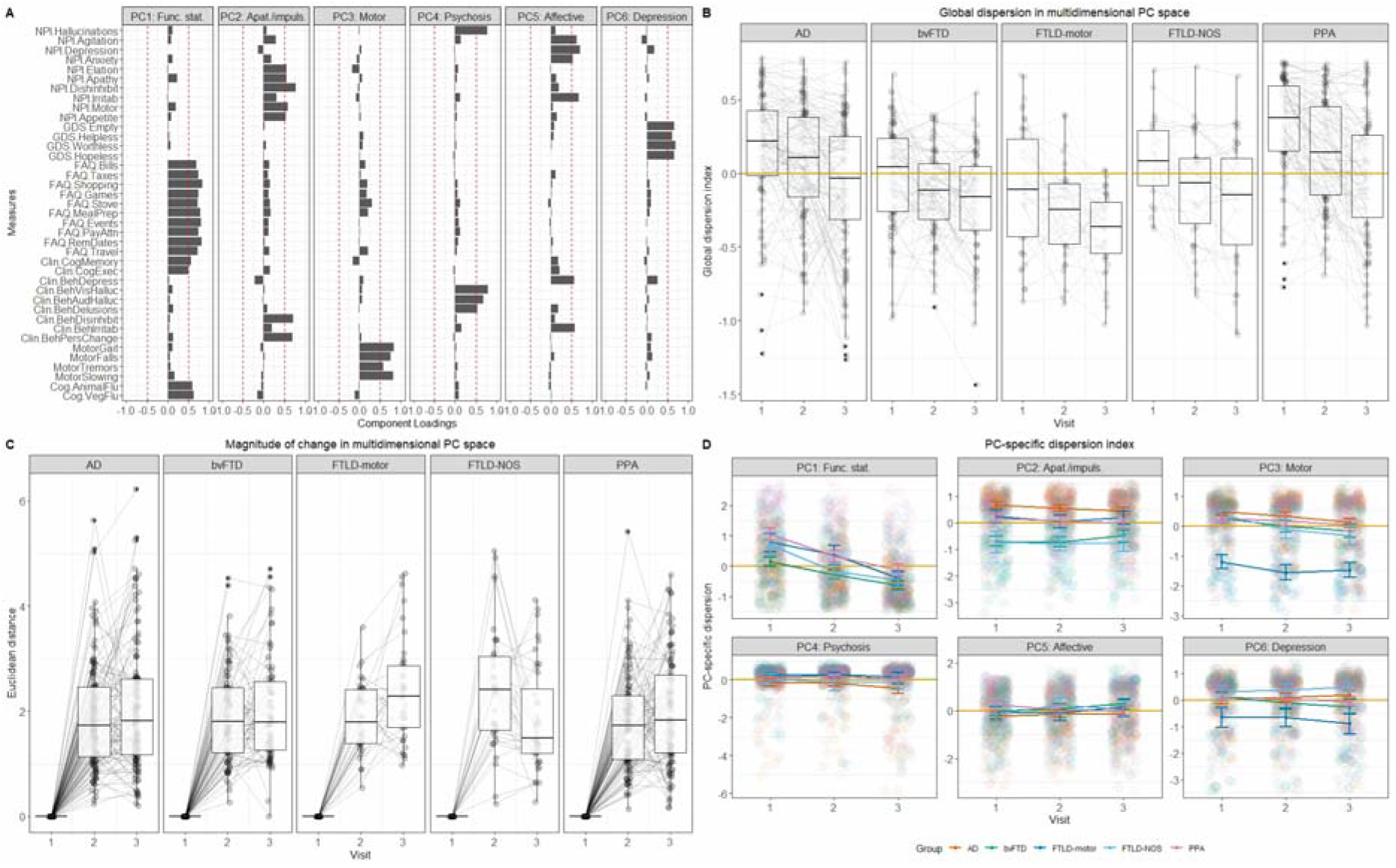
Axes, direction, nature, and magnitude of longitudinal phenotypic variation in patient groups within the multidimensional PC space. Panel **A)** Component loadings for clinical, cognitive and behavioural measures in the combined patient group (*N*=1,170, all time points included) on a varimax-rotated 6 component PCA solution. Panels indicate emerging components in the order of amount of overall variance explained. Red dotted lines represent component loading cut-offs (>|.5|). The 6 component solution explained 51% of the overall variance (PC1=19%, PC2=9%, PC3=7%, PC4=6%, PC5=5%, PC6=5%). Panel **B)** Global dispersion index where gold lines indicate the centroid of the multidimensional PC space. Convergence towards the centroid indicates progression of syndromes towards a de-differentiated pooled phenotype, whereas divergence from the centroid suggests that the syndrome differentiates from the others. Negative and positive values indicate direction of movement in the multidimensional PC space, relative to the centroid. For box-and-whisker plots, upper and lower bound of box correspond to upper and lower quartiles, black central line corresponds to median, and upper and lower end of whiskers correspond to maximum and minimum values, with data outside of whiskers indicating outliers. Panel **C)** Magnitude of phenotypic change for each individual in the multidimensional PC space calculated as the average Euclidean distance between their baseline and follow-up scores. Box-and-whisker characteristics same as aforementioned description. Panel **D)** Nature of PC-specific change in each group, with gold lines indicating the centroid of each PC. Error bars indicate standard error of the mean. PC=principal component; NPI=Neuropsychiatric Inventory; GDS=Geriatric Depression Scale; FAQ=Functional Activities Questionnaire; Clin=Clinical; Beh=Behavioural; Cog=Cognitive; Flu=Fluency; Func. Stat.=Functional Status; Apat./impuls.=Apathy/Impulsivity; AD=Alzheimer’s disease; bvFTD=behavioural variant frontotemporal dementia; FTLD=frontotemporal lobar degeneration; NOS=not otherwise specified; PPA=primary progressive aphasia.

### Some syndromes de-differentiate towards a pooled phenotype over time

Next, all PCs were considered together within a 6D “multidimensional PC space”, where each patient’s location is a proxy for their overall cognitive-behavioural profile at that given time point.

Using the global dispersion index, we first probed the direction of movement to ask “how phenotypically similar do neurodegenerative disorders become over time?” In all patient groups, the global dispersion index was correlated modestly (all *r*>.-22) to strongly (all *r*<-.58) with an independent disease severity measure (CDR-FTLD-SoB) (all *p*<.001; Supplementary Table 5), meaning that as individuals converged towards this de-differentiated average phenotype, their disease severity increased, assuring of its use as a data-driven proxy of phenotypic changes over time. Linear mixed-effect models fitted on the global dispersion index revealed all groups to change with time (*t*=−6.1; *p*<.001) with the largest time*group interactions in PPA (*t*=−2.4; *p*=.015). This pattern replicated when analyses were run including only individuals with stable diagnoses over time (Supplementary Results). On boxplots, PPA, as well as AD, gravitated towards the centroid (gold line) progressing towards a de-differentiated pooled phenotype, while bvFTD and FTLD-motor groups appeared to diverge to look different from this generic phenotype (Figure 1B).

Next, we asked “which groups show the greatest cognitive-behavioural changes over time?” (Figure 1C). Significant main effects for AD [*F*(2,371)=178.9; *p*<.001; η^2^=.49[.43-1]], bvFTD [*F*(2,263)=230; *p*<.001; η^2^=.64[.58-1]], FTLD-motor [*F*(2,113)=138.4; *p*<.001; η^2^=.71[.64-1]], FTLD-NOS [*F*(2,99)=51.1; *p*<.001; η^2^=.51[.39-1]] and PPA groups [*F*(2,309)=187.2; *p*<.001; η^2^=.55[.49-1]] emerged, with largest changes noted between baseline (Visit 1) and both follow-ups (all *p*<.001). Between-group comparisons at each time point revealed significant differences at the first follow-up [*F*(4,385)=2.9; *p*=.019; η =.03[0-1]] with FTLD-NOS group showing greater changes than PPA (*p*=.007) and AD (*p*=.029).

Third, we investigated the nature of phenotypic change in each group using linear mixed-effect models on PC-specific dispersion indices (Figure 1D). On PC1 (Functional status), largest changes over time were noted in PPA (*t*=−3.2; *p*=.001) and FTLD-motor (*t*=−2.2; *p*=.02) groups. On PC2 (Apathy/impulsivity), marked changes were noted in FTLD syndromes, including bvFTD (*t*=2.5; *p*=.01), FTLD-motor (*t*=2; *p*=.40) and FTLD-NOS groups (*t*=2.3; *p*=.019). For PC3 (Motor function), only main effects for Visit (*t*=−4.2; *p*<.001) and FTLD-motor group (*t*=−8; *p*<.001) were significant. On PC4 (Psychosis), a main effect of time was found (*t*=−2.5; *p*=.011) with no significant interactions. Turning to PC5 (Affective changes), the PPA group demonstrated the largest changes over time (*t*=−2.5; *p*=.010). Finally, on PC6 (Depression), the largest changes with time were found in the bvFTD (*t*=−3.2; *p*=.001) and FTLD-motor groups (*t*=−2.1; *p*=.031).

In summary, on inspection of the 6D multidimensional PC space, we found that AD and PPA tend to gravitate more towards a de-differentiated pooled phenotype over time. Across all syndromes, there are large phenotypic changes that occur from baseline, but the specific nature of this change depends on where each patient lies on the underlying dimensions present.

### Visualising phenotypic interdigitation and diversification in AD and FTLD

Using UMAP, we visualised and map longitudinal phenotypic interdigitation and diversification of AD/FTLD. In the “UMAP space”, inter-relation of distances and positions of data points provide a low-dimensional interpretable structure to longitudinal movement of patients.^57–60^ UMAP “snap-shots” of phenotypic convergence and divergence at the individual- and group-level are shown in Figures 2A-D. Animated plots displaying patient-specific trajectories of movement within the UMAP space can be viewed via this link: https://github.com/siddharthramanan/NACC_UMAP/tree/main/UMAP_animate.

**Figure 2.**
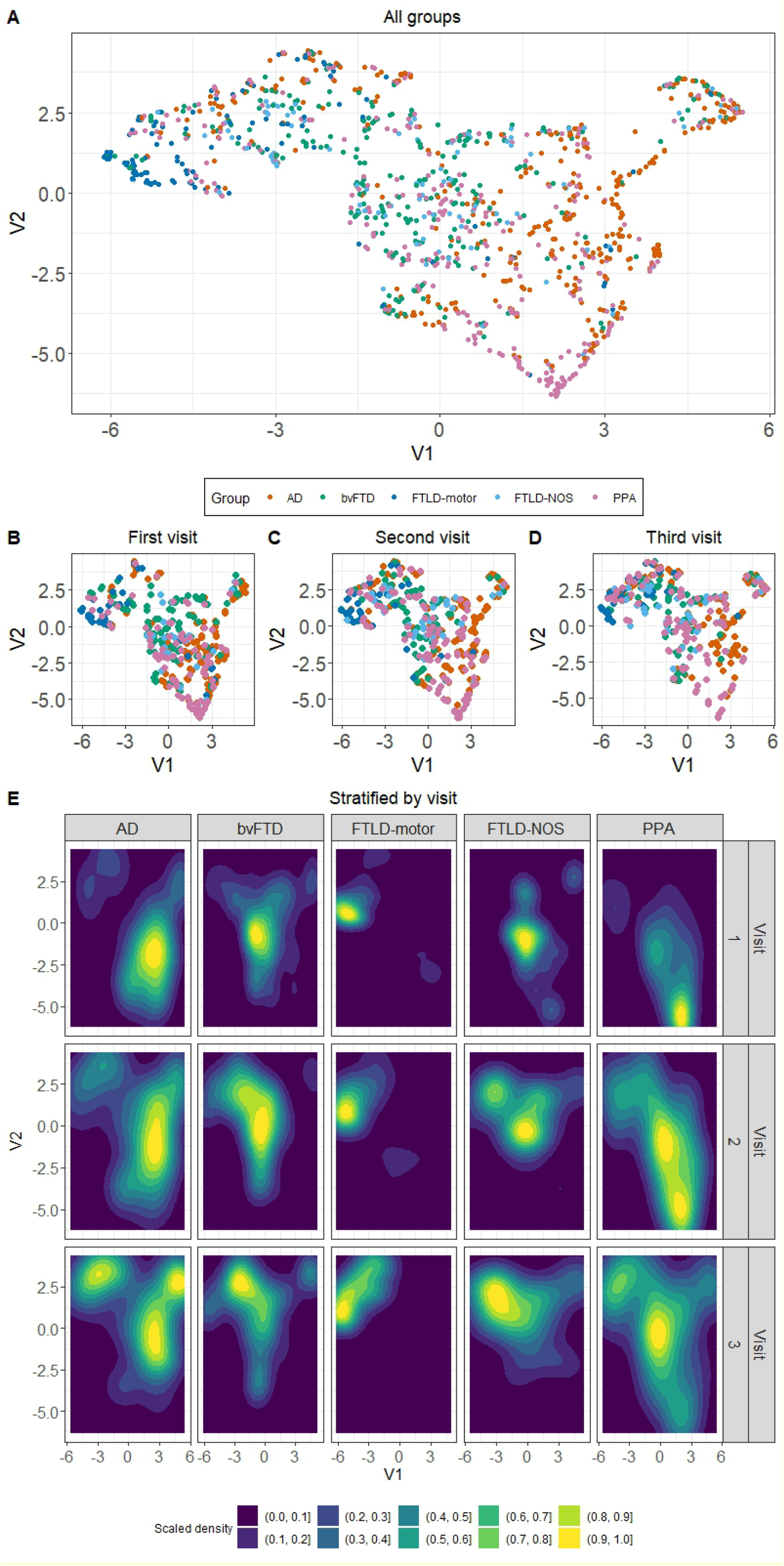
Low-dimensional UMAP embedding of patient groups. Panel **A)** UMAP embedding for all time points combined where V1 and V2 are the two UMAP axes. Panels **B, C, D)** UMAP embedding stratified by each visit. Panel **E)** Spatial kernel density estimation plots indicating the “movement” of each patient group within the UMAP space over time. Warm colours indicate the greatest density scaled to sample size. UMAP embedding initialised using the multidimensional PC space. UMAP=Uniform Manifold Approximation and Projection; PC=principal component; AD=Alzheimer’s disease; bvFTD=behavioural variant frontotemporal dementia; FTLD=frontotemporal lobar degeneration; NOS=not otherwise specified; PPA=primary progressive aphasia.

As a first step, we validated the statistical structure of the UMAP space to determine whether the embedding patterns were random or non-random (Supplementary Figure 9). The observed ANN values were far smaller than expected under the null hypothesis (observed ANN=.07 vs. mean null ANN=.125; *p*<.001), suggesting the UMAP embedding followed a non-random spatial distribution carrying information on graded group-level differences.

Next, we visualised the longitudinal movement patterns of each group in the UMAP space to understand dispersion vs. clustering over time (Figure 2E). All groups showed significant movement between baseline (Visit 1) and second follow-up (Visit 3) (all *z*>=1.8; all *p*<=.03; Supplementary Table 6) and this was most marked in the PPA group (baseline vs. first follow-up, *z*=7.2 and *p*<.001; baseline vs. second follow-up, *z*=13.3 and *p*<.001). Longitudinally within the PPA taxonomy, visualisations revealed that logopenic PPA patients moved into the AD space, semantic PPA moved to become tightly clustered in the bvFTD space, nonfluent PPA group either remained in the PPA space or gravitated to the FTLD-motor space intermingling with the mixed PPA group (patients originally labelled as “PPA”), and PPA-NOS gravitated into the FTLD-NOS space (Supplementary Figure 10). Taken together, there was striking and eventual visual convergence of many patients into the part of the UMAP space perennially occupied by the FTLD-motor group, with AD and PPA showing marked spatial diversification to spread across the UMAP space.

We then tested whether group-wise patterns of movement informed of convergence towards a de-differentiated pooled phenotype (de-differentiation hypothesis) or continuing differentiation (Supplementary Figure 11 and Supplementary Table 7). In these findings, clustering indicates homophily and within-group similarity, while overlap with the null suggests a profile de-differentiated from other syndromes (i.e., general phenotypic diversification). ANN findings were concordant with its counterpart global dispersion index, adding an important layer of spatiotemporal granularity. The AD group spatially clustered at baseline (*p*=.029) but dispersed over time to overlap with the null distribution (both *p*>.1) suggesting phenotypic divergence within a year from first assessment. BvFTD patients clustered at baseline and first follow-up (both *p*<.005) but diverged at second follow-up (Visit 3) to overlap with the null distribution (*p*>.1). In contrast, the FTLD-motor group continued to cluster at every time point, significantly different from the null hypothesis (all *p*<.005). The FTLD-NOS group was largely dispersed, although, at the first follow-up (Visit 2) there was some evidence for clustering compared to the null distribution (*p*<.005). The PPA group was dispersed at all time points, overlapping with the null (all *p*>.08), indicating inherent heterogeneity across assessments. Together, these analyses bring clear evidence for some groups (AD, bvFTD) to become less homogeneous with time, others (FTLD-motor) to remain homogeneous through longitudinal evaluations, and yet others (PPA, FTLD-NOS) to present heterogeneously across assessments.

### Linking AD-FTLD syndrome spaces to symptom prevalence and severity

Rank ordered performance for each PC within the UMAP space is displayed in Figure 3 (corresponding tertiles in Supplementary Figure 12). PC1 (Functional status) ranking contoured along the UMAP *y*-axis with the most impaired individuals in FTLD space. For PC2 (Apathy/impulsivity), the lowest ranks were again in the FTLD space, whereas for PC3 (Motor function), these were concentrated in the FTLD-motor space. Lowest performers on PC4 (Psychosis) were spread out and occupied by regions predominated by AD, PPA, and FTLD patients. PC5 (Affective changes) scores were lowest in the space occupied by individuals with FTLD syndromes. Finally, the lowest scorers on PC6 (Depression) were spread across the *y*-axis of the UMAP.

**Figure 3.**
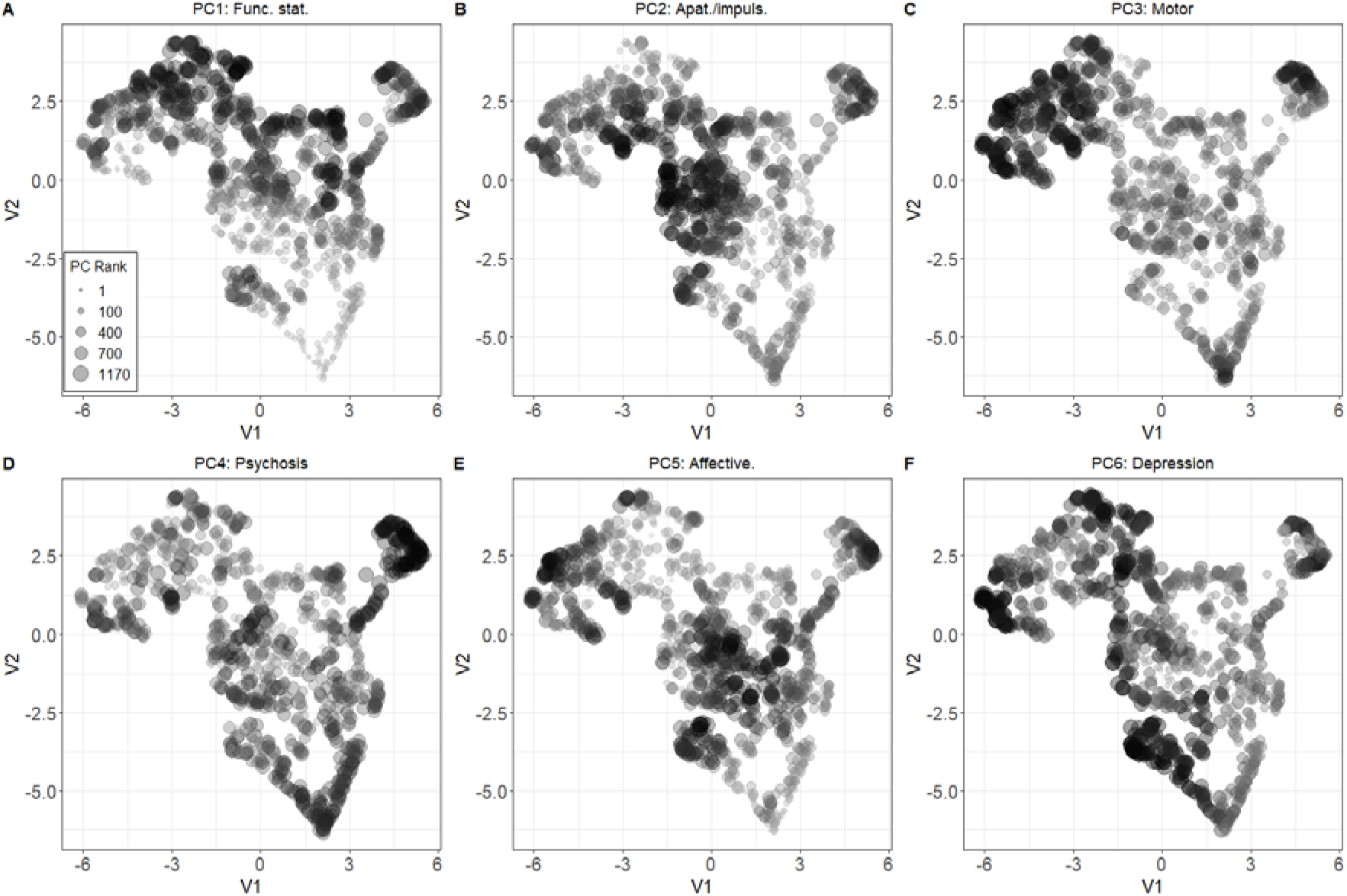
Rank dot plot for all PCs projected into the UMAP space. V1 and V2 are the two UMAP axes. Ranks are scaled by size. PC=principal component; UMAP=Uniform Manifold Approximation and Projection; Func. Stat.=Functional Status; Apat./impuls.=Apathy/Impulsivity.

Projecting the predominant symptom first recognised as a decline in a patient’s behaviour into the UMAP space, apathy was the most frequent transdiagnostic feature across time (Supplementary Figure 13). Disinhibition was most common in bvFTD and FTLD-motor groups, irritability in AD, and personality changes in bvFTD patients. Turning to cognitive changes, executive dysfunction was most common in FTLD syndromes, language changes in PPA and FTLD-motor conditions, and memory impairment in AD and bvFTD groups (Supplementary Figure 14).

### Predicting clinico-pathological endpoints from low-dimensional spaces

Focusing on survival, 100/209 patients (47%) died within 3 years of their first recorded NACC visit. Individuals who died within the first 3-years had significantly longer symptom duration (mean (SD): 5.4 (4.6) years) than those who died after the 3-year cutoff (mean (SD): 4.6 (2.9) years) and those who were not deceased (mean (SD): 4.2 (2.6) years) [*F*(2,384)=3.9; *p*=.019; η =.02[0-1]]. We then conducted three independent analyses exploring associations between survival status and cognitive-behavioural heterogeneity using global (UMAP coordinates and global dispersion scores) and local measures (PC-specific scores). For global metrics, we found no significant associations between survival at 36 months and baseline location in the UMAP space with linear models (*z*=.3, *p*=.7; main effects: all *z*<.3, *p*>.1) or non-linear spatial regressions (*z*=−.6, *p*>.1). Examining this association in the multidimensional PC space, we found no significant statistical associations (all *z* <|.5|, all *p*>.1), even when models were run within individual patient groups.

Focusing on local indices, in AD, survival at 36 months was significantly predicted by starting level of impairment on PC1 (Functional status; *z*=−1.9, *p*=.04) and PC4 (Psychosis; *z*=−2, *p*=.04) (Supplementary Table 8). In FTLD-motor patients, starting level of impairment on PC1 (Functional status; *z*=−2.3, *p*=.01), and starting point and travelled distances on PC3 (Motor function; starting point *z*=2.5, *p*=.01; travelled distance *z*=−2.2, *p*=.02) significantly predicted survival at 36 months. In PPA, the starting level of impairment on PC5 (Affective changes; *z*=2.6, *p*=.008) and travelled distances on PC1 (Functional status; *z*=−2.4, *p*=.01) and PC3 (Motor function; *z*=−2.2, *p*=.02) significantly predicted survival at 36 months. No significant associations emerged in the bvFTD group and model convergence failed in FTLD-NOS due to small numbers. These findings suggest baseline levels and accrual of PC-specific deficits, rather than global cognitive-behavioural performance, differentially predict survival in AD, FTLD-motor and PPA syndromes.

Turning to pathology, we found evidence for spatial clustering of pathological groups significantly differently from the null in AD pathology, Corticobasal Degeneration (CBD), FTLD-Tau, LBD, PSP pathology (all *p*<.001), FTLD-Pick (*p*=.019), FTLD-Tar DNA binding protein-43 (TDP-43; *p*=.002) and Other pathological groups (*p*=.015) (Figure 4).

**Figure 4.**
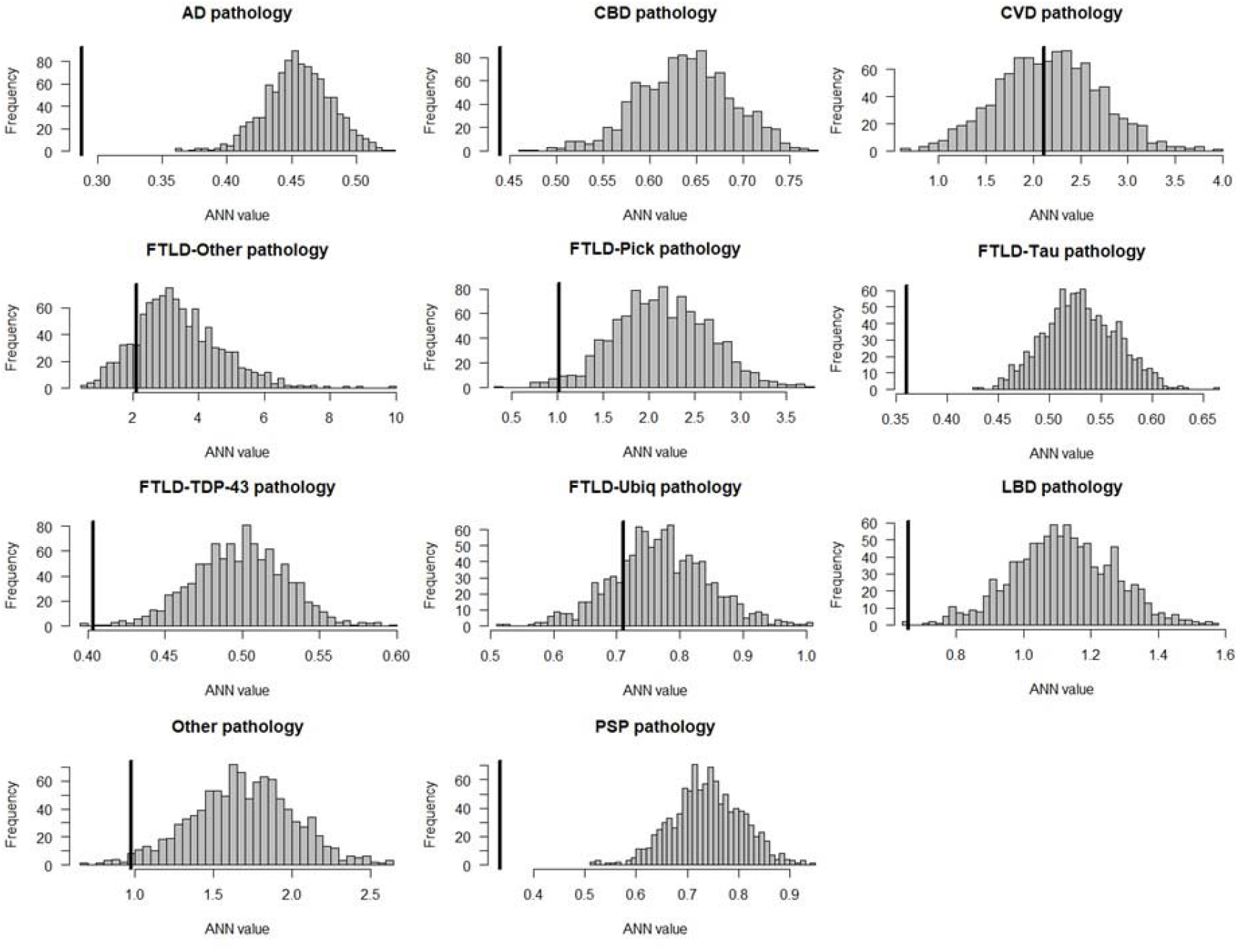
Average Nearest Neighbours analyses testing the statistical structure of group-wise embedding in the UMAP space in each pathological group. Histogram of null distribution of average nearest neighbours analysis compared to our observed average nearest neighbour value from the UMAP embedding (black bar). ANN=average nearest neighbours; AD=Alzheimer’s disease; FTLD=frontotemporal lobar degeneration; TDP-43=Tar DNA binding protein-43; LBD=Lewy Body Disease; CBD=Corticobasal Degeneration; CVD=cardiovascular disease; Ubiq=ubiquitin; PSP=Progressive Supranuclear Palsy; UMAP=Uniform Manifold Approximation and Projection.

These groups displayed non-random, local spatial clustering patterns (Figure 5A-B); however, we found no clear one-to-one mapping between clinical and pathological labels, with the exception of pathological PSP label (corresponding largely to FTLD-motor clinical label) and FTLD-Pick pathology (corresponding to either bvFTD or PPA clinical groups).

**Figure 5.**
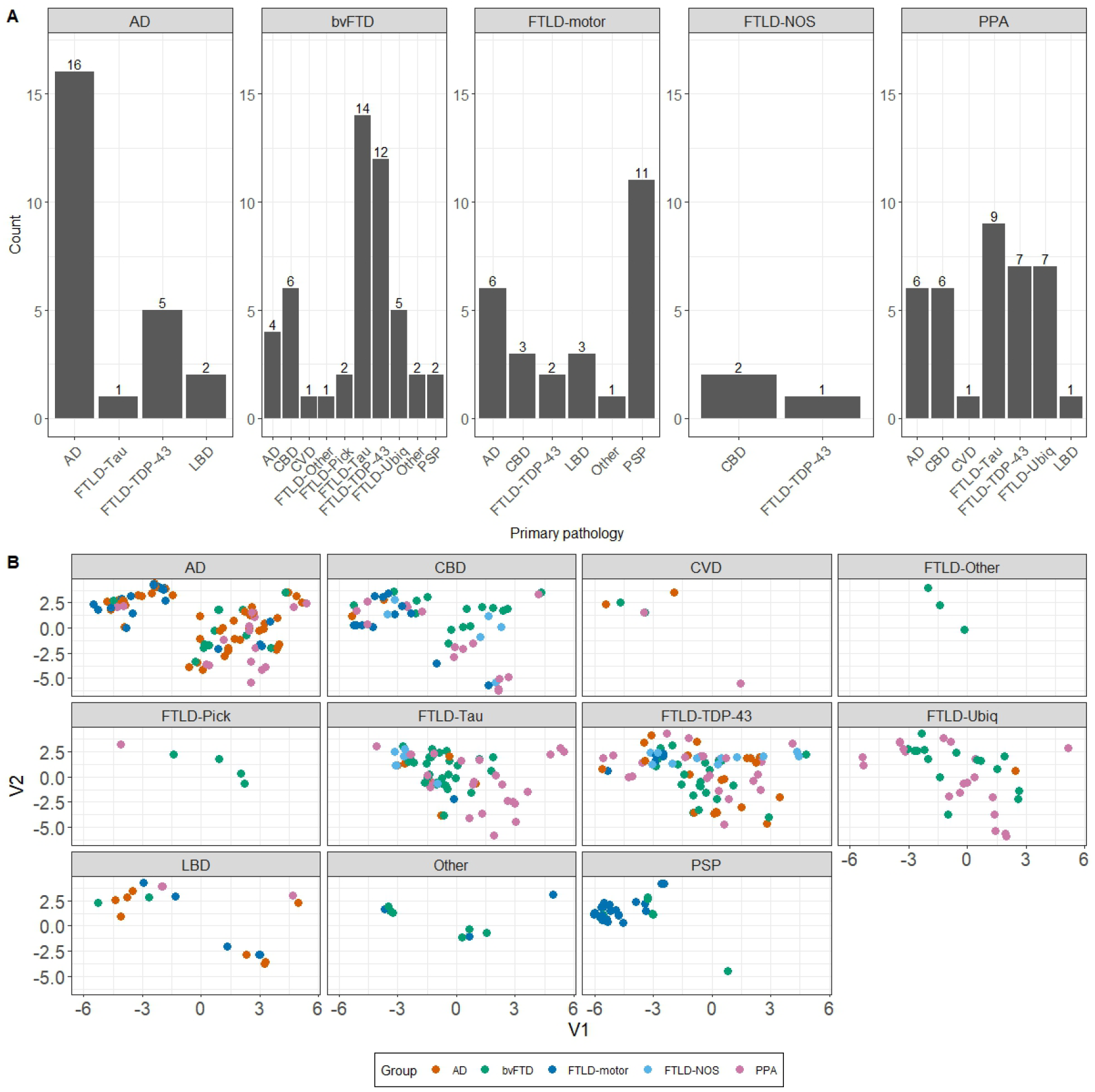
Frequency count and UMAP locations for pathological diagnoses. Panel **A)** frequency counts where facets display the five clinical diagnoses considered here with *x*-axis displaying the major pathological diagnoses reported in each clinical group; Panel **B)** clinicopathological mapping of AD and FTLD patients in the UMAP space faceted by pathological groups. Facets display 11 primary pathological brackets under which patients were classified. AD=Alzheimer’s disease; FTLD=frontotemporal lobar degeneration; TDP-43=Tar DNA binding protein-43; LBD=Lewy Body Disease; CBD=Corticobasal Degeneration; CVD=cerebrovascular disease; Ubiq=ubiquitin; PSP=Progressive Supranuclear Palsy; bvFTD=behavioural variant frontotemporal dementia; FTLD=frontotemporal lobar degeneration; NOS=not otherwise specified; PPA=primary progressive aphasia.

All clinical syndromes located across multiple pathologies (Figure 6). No significant differences from the null hypothesis were noted for pathological groups of CVD (*p*=.45), FTLD-Other (*p*=.15), and FTLD-Ubiquitin (*p*=.2) suggesting random spatial location in the UMAP; however, extremely small sample sizes in CVD and FTLD-Other may underpin these results.

**Figure 6.**
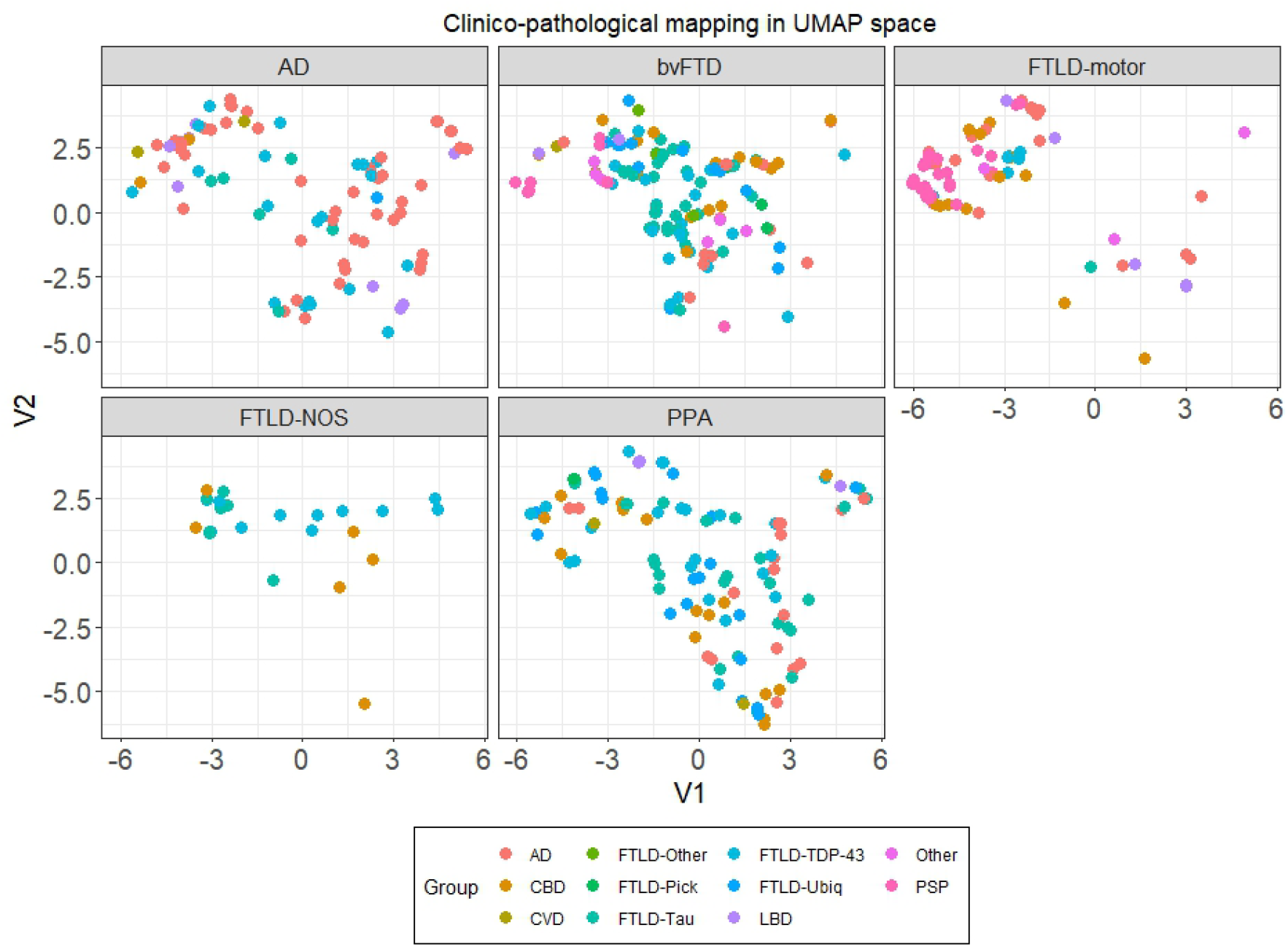
Clinicopathological mapping of AD and FTLD patients in the UMAP space faceted by clinical groups. Each panel represents a clinical group. AD=Alzheimer’s disease; FTLD=frontotemporal lobar degeneration; TDP-43=Tar DNA binding protein-43; LBD=Lewy Body Disease; CBD=Corticobasal Degeneration; CVD=cardiovascular disease; Ubiq=ubiquitin; PSP=Progressive Supranuclear Palsy; bvFTD=behavioural variant frontotemporal dementia; FTLD=frontotemporal lobar degeneration; NOS=not otherwise specified; PPA=primary progressive aphasia.

Finally, we examined whether pathological labels predicted PC-specific decline. For PC1 (Functional Status), no significant interactions emerged, however, significant main effects of time (Visit 2: *t*=−2.2; *p*=.025; Visit 3: *t*=−3.9; *p*=.001) and group for pathological labels of CBD (*t*=3.2; *p*=.001), FTLD TDP-43 (*t*=2.1; *p*=.03), FTLD-Ubiquitin (*t*=2.4; *p*=.01), and PSP (*t*=2.2; *p*=.023) were noted. For PC2 (Apathy/impulsivity), the FTLD-Other pathology group displayed significant performance changes at the third visit (*t*=2.3; *p*=.02). For PC3 (Motor function), a significant main effect for the PSP pathology group was noted (*t*=−5.9; *p*<.001), however, no interaction terms emerged to be significant. For PC4 (Psychosis), largest performance changes over time were noted in LBD pathology patients (Visit 2: *t*=2.2; *p*=.02; Visit 3: *t*=3.4; *p*<.001), and at the final visit for patients with pathological labels of FTLD TDP-43 (*t*=2.8; *p*=.004) and PSP (*t*=2.6; *p*=.008). For PC5 (Affective changes), a significant main effect emerged for the FTLD-Ubiquitin pathology group (*t*=2.2; *p*=.02). For PC6 (Depression), patients with CBD pathology displayed marked performance changes between first and second visit (*t*=−2.2; *p*=.02).

Together, the findings show that despite evidence for the de-differentiation hypothesis, the extent to which this occurs globally across diagnoses itself does not seem to predict mortality at 3 years following baseline. Instead, Functional Status (PC1 score) is a significant predictor of survival across most diagnoses, with other PCs holding predictive power for survival in a diagnosis-specific manner, such as psychosis in AD, motor function in FTLD and affective status in PPA. While pathological groups do cluster within the UMAP space, there does not appear to be a clear one-to-one mapping between pathology and outcomes, and no strong predictive power of pathological substrates for cognitive-behavioural devolution, with the exception of associations between apathy/impulsivity (PC2) and FTLD-Other pathologies, psychosis (PC4) and LBD, and depression (PC6) and CBD pathology.

## Discussion

Untangling the graded, dynamic and longitudinal phenotypic variations in AD/FTLD has remained a continuing clinical and analytical challenge, with implications for diagnosis, management, and prognostication.^66^ To address this issue, we combined three scientific pillars to revisit longitudinal clinical variations through a new lens: large longitudinal data sampling a range of AD/FTLD presentations and symptoms, advanced analytics, and graded multidimensional geometries. The structured longitudinal variability in AD/FTLD, as revealed by our PCA, was underpinned by six fundamental dimensions (approximating Functional changes, Apathy/impulsivity, Motor function, Psychosis, Affective changes, and Depression). Most patients did not demonstrate features along a single axis; rather, clinical groups located in graded multidimensionally-defined subregions of the space. Dimensions were inherently transdiagnostic in nature. At this stage, a clear advantage of using PCA as an analytic step was that dimensions emerged as linear combinations of input variables; therefore, they could be readily back-translated to clinical features. Specifically, we could locate and inter-relate individuals on each dimension, knowing well what specific measures likely predicated this phenotypic variation. Further, by selecting key tests loading heavily on particular PCs as assessment proxies for those dimensions, we could understand which key measures held potential to be most sensitive to transdiagnostic phenotypic variations in neurodegeneration. This understanding could inform future steps such as improving tests for neuropsychological assessment and reducing the length of test batteries.^67^ However, a subsequent challenge that emerged was the difficulty associated with interpreting changes along all six PCs in parallel. We therefore complemented PCA with strengths of advanced neighbour embedding methods, namely UMAP. Through the UMAP space, we reinforced the key idea that, with the exception of FTLD-motor syndromes, separate AD/FTLD variants do not tightly or consistently cluster over time. They display high within as well as between-group variance, magnifying with time. Thus, some individuals diversify to resemble different clinical groups while others remain true-to-type, irrespective of their original diagnostic label.

Projecting well-described clinical labels into our multidimensional PCA and UMAP spaces and linking to clinico-pathological endpoints added important layers of clinical interpretability. For example, AD patients exhibited homophily (i.e., self-similarity) at baseline but progressed over time to resemble other neurodegenerative syndromes. Longitudinal dispersion of AD cases into specific parts of the UMAP space (which were occupied by PPA and FTLD patients) was not predicated by their pathological label; rather, it reflected increasing psychosis and functional decline – two variables that further predicted their survival status at 3 years. These findings form an important visualisable addition to the literature of AD cognitive-behavioural progression dynamics, in line with extant evidence for increasing disease severity and accrual of additional cognitive complaints to contribute to phenotypic resemblance of AD with other dementia syndromes.^68,69^ Turning to PPA, these patients displayed inherent heterogeneity and phenotypic de-differentiation at every time point. The visual movement of well-described PPA nosological entities matched previously described patterns,^70^ where logopenic PPA moved into the AD space, semantic PPA progressed to resemble bvFTD, and nonfluent PPA mirrored FTLD-motor syndromes over time. Importantly, the heterogeneity in this group largely arises from a number of intermediate patients, who have been poorly described in the literature yet have often formed the majority of the PPA group. These individuals progressed over time to straddle the AD and FTLD syndrome spaces. PPA patients further went on to accumulate functional and affective deficits with time,^71^ and the accrual of functional and motor impairments predicted their survival status at 3 years post-baseline.^72^ In the context of ongoing debates, the current findings concur with a growing literature in PPA supporting the emergence of non-linguistic and functional changes which (i) are sometimes notable in early stages of the disease, when assessed appropriately;^73–75^ (ii) are present systematically in most variants, irrespective of the type and severity of their language disorder;^30,32,76^ and (iii) may not directly relate to specific pathological substrates.^77^ As functional decline and motor changes hold prognostic importance in predicting mortality in PPA and other neurodegenerative syndromes,^78^ these features must be considered as important targets of symptomatic and underlying disease interventions in these syndromes.

In contrast to phenotypic de-differentiation in AD and PPA, FTLD syndromes did not converge towards a de-differentiated pooled phenotype, instead displayed magnification of specific cognitive-behavioural changes.^79^ For example, apathy/impulsivity emerged as a longitudinal transdiagnostic marker of all FTLD syndromes, as has been noted before.^80^ Depression was prominent in bvFTD and FTLD-motor groups, and functional changes in FTLD-motor patients. FTLD syndromes can be extremely challenging to diagnose, with recent recommendations for the principled search and adoption of phenotypic markers, such as apathy and socioemotional cognition (closely reflecting specific micro-/macro-level neural network alterations) that could help in its accurate diagnosis.^81,82^ Profound neuropsychiatric changes, amidst relatively preserved cognitive performance, can be noted in the earliest stages of some FTD syndromes.^83^ Our magnification results extend this finding to show that features such as apathy, depression and functional changes significantly magnify with time in FTLD syndromes, suggesting the importance of examining the value of these markers in predicting diagnosis at early stages and predicting important clinical end-points such as institutionalisation and death. In the UMAP space, the locations of different FTLD variants appeared to reflect their relative rates of phenotypic diversification. For example, on the one end of the space lay FTLD-motor cases, who displayed homophily, clustered tightly over time and largely continued to have motor symptoms. In another part of the space were FTLD-NOS, longitudinally interdigitating with FTLD, PPA and AD variants. BvFTD were somewhere midway, progressing to show features of other syndromes but only by their third annual assessment, suggesting relatively faster phenotypic diversification in this group as compared to motor variants of FTLD. More generally, the broader FTLD space was uniformly underpinned by marked apathy/impulsivity, disinhibition, executive dysfunction, psychosis and affective disturbances, all of which are increasingly suggested to be transdiagnostic features closely tied to degeneration of frontal and striatal brain circuitry noted across FTLD variants.^84,85^ Motor dysfunction on the other hand, was largely restricted to the quadrant of the UMAP occupied mainly by FTLD-motor patients, indicating the segregation of this symptom and its resulting phenotypes from other FTLD variants. Furthermore, longitudinal accrual of motor symptoms, along with functional changes, closely predicted survival in FTLD-motor patients.^78^ Apathy/impulsivity, despite its established prognostic importance in FTLD,^86^ did not emerge as a significant predictor of mortality in these clinical data, potentially due to floor effects and thus a lack of variance. Turning to symptom-syndrome-pathology mapping in FTLD, we found relatively specific links only between PSP pathology and a clinical diagnosis of FTLD-motor conditions. Symptoms such as apathy in patients with FTLD-Other pathology and psychosis in FTLD-TDP-43 and PSP pathological groups magnified three years post-baseline assessment, while depression in those with CBD pathologies accentuated within two years post-baseline.^84^ All other FTLD pathologies were found to be present across a range of clinical diagnoses,^87^ including AD and PPA. Overall, the geometries revealed that, despite the presence of different pathologies and a complex relationship between symptoms and underlying pathological contributors, many FTLD cases became more alike over time.^16^ An important implication of this finding is the need for better trans-syndromic functional markers of molecular pathologies, as instantiated at both the macro- and micro-circuit level.^85,88^ Specifically, it is the conjunction of pathological protein changes, neurotransmitter and cell-level aberrations, and intrinsic structural/functional network-level breakdowns that, when modelled with a coherent framework, could shed light on the specific mechanisms underpinning phenotypic signatures of neurodegenerative syndromes. It is, therefore, important for future work to complement transdiagnostic phenotypic studies with micro/macro-level circuit modelling, via structural and functional neuroimaging tools, to unravel the “molecular nexopathies” that contribute to phenotypic diversity.^88,89^

This study had a number of limitations that warrant discussion. The power of large samples from multi-centre retrospective data is tempered by missing data or missing available measures and samples. The historic focus of the NACC database has been the AD pathological continuum. Therefore, issues arise with regards to the presence of sufficient non-AD cases and sampling of rare, atypical presentations of AD/FTLD (e.g., dysexecutive AD, behavioural variant AD, Posterior Cortical Atrophy, PPA with apraxia of speech). Moreover, a significant proportion of our study sample that came to post-mortem had an advanced disease duration and may have biased our de-differentiated phenotypic analyses. It is, therefore, important for future work to sample individuals at earlier disease stages. Like with any large multi-centre consortium dataset, there are also limitations of missing data and assessments. The current study aimed to address some of these issues by selecting a sample that was representative of the wider AD/FTLD typical, atypical and intermediate phenotypic space, yet had minimal missing data (5.9% missing data in the current study). However, in an effort to balance missing data across available assessments, a number of measures that are important to expand on our understanding of FTLD phenotypic presentations (e.g., detailed language, social cognitive, and other multi-domain neuropsychological assessments) were either not available/complete or could not be included in the current study. As a result, the output of our PCA and UMAP analyses and their capacity for phenotype differentiation are limited to the measures included in the current study. Specifically, the inclusion of AD, FTLD and PPA groups, assessed over multiple time points, resulted in a preponderance of neuropsychiatric and functional assessments, over cognitive and language tests, that were common and uniformly available across all individuals. This pattern of data availability has implications to future work examining longitudinal phenotyping in AD/FTLD, where (i) the use of the NACC dataset in FTD and PPA may be more suitable to examine functional and neuropsychiatric markers of disease presentation and evolution, and (ii) certain major syndromes of high diagnostic specificity (such as primary progressive apraxia of speech) may be represented to a limited extent and/or have limited data in the NACC dataset. Future studies sampling a larger symptom/syndrome space with a broader battery of multi-domain cognitive-behavioural tests will be important to draw out greater variations across multiple neurodegenerative syndromes. While the longitudinal element was a novel analysis in this study, the dispersion metrics over three years may not be sensitive to slowly progressive conditions such as semantic variant PPA.^90^ Furthermore, we benefitted from being able to link clinical, cognitive and pathological levels of heterogeneity; however, additional data on fluid biomarkers, brain changes, and genetic mutations will be the key to understanding other disease mechanisms underpinning the emergence of clinical heterogeneity in these syndromes. While the current study did benefit from the addition of pathological data, we were only able to consider contributions of singular pathologies in explaining phenotypic changes. It is important for future work to account for mixed/concurrent neurodegenerative and vascular pathologies, and their relative contributions to clinical presentation, progression and end-points such as survival. Extension of our framework to presymptomatic patients and Mild Cognitive Impairment, will be pivotal to revealing the emergence and evolution of clinical heterogeneity in the pathological ageing spectrum.

This work has important clinical and research implications, some of which are listed here. First, the decision to ‘lump’ or ‘split’ disorders into subtypes may advance disease nosology. Yet, without insights into common mechanisms and systematic graded variations cutting across disease boundaries, we cannot develop, validate and apply disease-modifying treatments benefitting the entire clinical dementia spectrum. Second and relatedly, embracing the systematic patient variations within a coherent multidimensional space offers the chance to explain why many individual patients do not confirm perfectly to paradigmatic exemplars of categorical labels and why these labels change between clinic visits over time. Third, it is also important to recognise the dynamic evolution of progressive degenerative conditions, and that such dynamism is a major feature of all progressive disorders, not simply unique to rare conditions. In turn, the gradual de-differentiation over time is consistent with current efforts to capture and diagnose neurodegenerative syndromes at earlier stages, which in turn could help with predicting possible trajectories of phenotypic diversification. Fourth, understanding this longitudinal dynamism is also important for patients and families. Specifically, in the earliest stages of disease or at initial diagnosis, an understanding of this phenotypic diversification and longitudinal dynamism could help in explaining what clinical features the patients and families may observe as time goes by, and help to leverage the early emergence of some features in future prognostic decision making, such as the relationship between the emergence of apathy and early death in FTLD syndromes.^86^ Fifth, by constructing multidimensional phenotypic geometries, our approach further elucidates the transdiagnostic pathogenesis of cognitive-behavioural heterogeneity to accommodate a range of clinical characterisations and permit a granular examination of graded within/between-group variations.^91^ This variation, is sometimes treated as ‘noise’ in a diagnostic category, thus precluding observation of effects in category-based analyses. However, these variations can be systematic and carry important information on individual-level mechanisms of phenotypic diversification, over-and-above what is explained by over-general labels such as “atypical presentation” or categorical stages of disease progression (e.g., mild vs. severe disease). Sixth, embracing the variation in a low-dimensional phenotypic landscape enables new approaches to precision medicine and personalised care planning rather than Procrustean management. Accordingly, the key axes of phenotypic variation also offer an opportunity to revise and update training material for clinicians outside of neurodegenerative specialist centres. Seventh, decoding the structure and dynamics of such variation within a manifold potentially supports new descriptive research frameworks of the architecture of phenotypic heterogeneity in dementia syndromes,^59^ in turn, informing design of targeted diagnostic markers and symptomatic treatments potentially titrated to reflect inter-individual positioning on dimensions, irrespective of clinical category. This holds significant potential to provide tailored disease-management information for clinicians, patients, families, and carers.

## Open access

For the purpose of open access, the authors have applied a CC BY public copyright licence to any Author Accepted Manuscript version arising from this submission.

## Supporting information

UMAP Plotly

## Data Availability

The National Alzheimers Coordinating Center dataset are freely available through request on their official website (https://naccdata.org/).

## Acknowledgements

The NACC database is funded by NIA/NIH Grant U24 AG072122. NACC data are contributed by the NIA-funded ADRCs: P30 AG062429 (PI James Brewer, MD, PhD), P30 AG066468 (PI Oscar Lopez, MD), P30 AG062421 (PI Bradley Hyman, MD, PhD), P30 AG066509 (PI Thomas Grabowski, MD), P30 AG066514 (PI Mary Sano, PhD), P30 AG066530 (PI Helena Chui, MD), P30 AG066507 (PI Marilyn Albert, PhD), P30 AG066444 (PI John Morris, MD), P30 AG066518 (PI Jeffrey Kaye, MD), P30 AG066512 (PI Thomas Wisniewski, MD), P30 AG066462 (PI Scott Small, MD), P30 AG072979 (PI David Wolk, MD), P30 AG072972 (PI Charles DeCarli, MD), P30 AG072976 (PI Andrew Saykin, PsyD), P30 AG072975 (PI David Bennett, MD), P30 AG072978 (PI Neil Kowall, MD), P30 AG072977 (PI Robert Vassar, PhD), P30 AG066519 (PI Frank LaFerla, PhD), P30 AG062677 (PI Ronald Petersen, MD, PhD), P30 AG079280 (PI Eric Reiman, MD), P30 AG062422 (PI Gil Rabinovici, MD), P30 AG066511 (PI Allan Levey, MD, PhD), P30 AG072946 (PI Linda Van Eldik, PhD), P30 AG062715 (PI Sanjay Asthana, MD, FRCP), P30 AG072973 (PI Russell Swerdlow, MD), P30 AG066506 (PI Todd Golde, MD, PhD), P30 AG066508 (PI Stephen Strittmatter, MD, PhD), P30 AG066515 (PI Victor Henderson, MD, MS), P30 AG072947 (PI Suzanne Craft, PhD), P30 AG072931 (PI Henry Paulson, MD, PhD), P30 AG066546 (PI Sudha Seshadri, MD), P20 AG068024 (PI Erik Roberson, MD, PhD), P20 AG068053 (PI Justin Miller, PhD), P20 AG068077 (PI Gary Rosenberg, MD), P20 AG068082 (PI Angela Jefferson, PhD), P30 AG072958 (PI Heather Whitson, MD), P30 AG072959 (PI James Leverenz, MD), ARTFL Consortium (U54 NS092089; MPIs Adam Boxer, MD PhD and Howard Rosen, MD), the ALLFTD Consortium (U19: AG063911; MPIs Bradley Boeve, MD, Adam Boxer, MD PhD and Howard Rosen, MD), the UPenn FTD Center (PI Murray Grossman, MD), and the Northwestern University PPA Program (R01 AG077444; PI M-Marsel Mesulam, MD).

## References

1. Au R, Piers RJ, Lancashire L. Back to the future: Alzheimer’s disease heterogeneity revisited. Alzheimer’s & Dementia: Diagnosis, Assessment & Disease Monitoring. 2015;1(3):368.

2. McKhann GM, Knopman DS, Chertkow H, et al. The diagnosis of dementia due to Alzheimer’s disease: Recommendations from the National Institute on Aging-Alzheimer’s Association workgroups on diagnostic guidelines for Alzheimer’s disease. Alzheimer’s & dementia. 2011;7(3):263–269.

3. Gorno-Tempini ML, Hillis AE, Weintraub S, et al. Classification of primary progressive aphasia and its variants. Neurology. 2011;76(11):1006–1014.

4. Armstrong MJ, Litvan I, Lang AE, et al. Criteria for the diagnosis of corticobasal degeneration. Neurology. 2013;80(5):496–503.

5. Höglinger GU, Respondek G, Stamelou M, et al. Clinical diagnosis of progressive supranuclear palsy: the movement disorder society criteria. Movement Disorders. 2017;32(6):853–864.

6. Snowden J, Goulding PJ, David N. Semantic dementia: a form of circumscribed cerebral atrophy. Behavioural Neurology. 1989;2(3):167–182.

7. Hodges JR, Patterson K. Semantic dementia: a unique clinicopathological syndrome. Lancet Neurol. 2007;6(11):1004–1014.

8. Rascovsky K, Hodges JR, Knopman D, et al. Sensitivity of revised diagnostic criteria for the behavioural variant of frontotemporal dementia. Brain. 2011;134(9):2456–2477.

9. Sajjadi SA, Patterson K, Arnold RJ, Watson PC, Nestor PJ. Primary progressive aphasia: a tale of two syndromes and the rest. Neurology. 2012;78(21):1670–1677.

10. Ramanan S, Roquet D, Goldberg ZL, et al. Establishing two principal dimensions of cognitive variation in logopenic progressive aphasia. Brain communications. 2020;2(2):fcaa125.

11. Tractenberg RE, Aisen PS, Weiner MF, Cummings JL, Hancock GR. Independent contributions of neural and “higher-order” deficits to symptoms in Alzheimer’s disease: a latent variable modeling approach. Alzheimer’s & Dementia. 2006;2(4):303–313.

12. Alladi S, Xuereb J, Bak T, et al. Focal cortical presentations of Alzheimer’s disease. Brain. 2007;130(Pt 10):2636–2645.

13. Ramanan S, El-Omar H, Roquet D, et al. Mapping behavioural, cognitive and affective transdiagnostic dimensions in frontotemporal dementia. Brain Communications. 2023;5(1):fcac344.

14. Lambon Ralph MA, Patterson K, Graham N, Dawson K, Hodges JR. Homogeneity and heterogeneity in mild cognitive impairment and Alzheimer’s disease: a cross-sectional and longitudinal study of 55 cases. Brain. 2003;126(Pt 11):2350–2362.

15. Roy ARK, Datta S, Hardy E, et al. Behavioural subphenotypes and their anatomic correlates in neurodegenerative disease. Brain Commun. 2023;5(2):fcad038.

16. Murley AG, Coyle-Gilchrist I, Rouse MA, et al. Redefining the multidimensional clinical phenotypes of frontotemporal lobar degeneration syndromes. Brain. 2020;143(5):1555–1571.

17. Brenowitz WD, Hubbard RA, Keene CD, et al. Mixed neuropathologies and estimated rates of clinical progression in a large autopsy sample. Alzheimer’s & Dementia. 2017;13(6):654–662.

18. Josephs KA, Hodges JR, Snowden JS, et al. Neuropathological background of phenotypical variability in frontotemporal dementia. Acta Neuropathologica. 2011;122(2):137–153.

19. Mehta RI, Schneider JA. What is ‘Alzheimer’s disease’? The neuropathological heterogeneity of clinically defined Alzheimer’s dementia. Current opinion in neurology. 2021;34(2):237–245.

20. Logroscino G, Capozzo R, Tortelli R, Marin B. Current Issues in Randomized Clinical Trials of Neurodegenerative Disorders at Enrolment and Reporting: Diagnosis, Recruitment, Representativeness of Patients, Ethnicity, and Quality of Reporting. In: The Right Therapy for Neurological Disorders. 2016:24–36.

21. Bradford A, Kunik ME, Schulz P, Williams SP, Singh H. Missed and delayed diagnosis of dementia in primary care: prevalence and contributing factors. Alzheimer Disease & Associated Disorders. 2009;23(4):306–314.

22. Jutten RJ, Sikkes SA, Van der Flier WM, et al. Finding treatment effects in Alzheimer trials in the face of disease progression heterogeneity. Neurology. 2021;96(22):e2673–e2684.

23. Ryan J, Fransquet P, Wrigglesworth J, Lacaze P. Phenotypic heterogeneity in dementia: a challenge for epidemiology and biomarker studies. Frontiers in public health. 2018;6:181.

24. Husain M. Transdiagnostic neurology: neuropsychiatric symptoms in neurodegenerative diseases. Brain. 2017;140(6):1535–1536.

25. Insel T, Cuthbert B, Garvey M, et al. Research domain criteria (RDoC): toward a new classification framework for research on mental disorders. In. Vol 167: Am Psychiatric Assoc; 2010:748–751.

26. Astle DE, Holmes J, Kievit R, Gathercole SE. Annual Research Review: The transdiagnostic revolution in neurodevelopmental disorders. Journal of Child Psychology and Psychiatry. 2022;63(4):397–417.

27. Lombardo MV, Lai M-C, Baron-Cohen S. Big data approaches to decomposing heterogeneity across the autism spectrum. Molecular Psychiatry. 2019;24(10):1435–1450.

28. Halai AD, Woollams AM, Lambon Ralph MA. Using principal component analysis to capture individual differences within a unified neuropsychological model of chronic post-stroke aphasia: Revealing the unique neural correlates of speech fluency, phonology and semantics. Cortex. 2017;86:275–289.

29. Ingram RU, Halai AD, Pobric G, Sajjadi S, Patterson K, Lambon Ralph MA. Graded, multidimensional intra- and intergroup variations in primary progressive aphasia and post-stroke aphasia. Brain. 2020;143(10):3121–3135.

30. Ramanan S, Irish M, Patterson K, Rowe JB, Gorno-Tempini ML, Lambon Ralph MA. Understanding the multidimensional cognitive deficits of logopenic variant primary progressive aphasia. Brain. 2022;145(9):2955–2966.

31. Jones D, Lowe V, Graff-Radford J, et al. A computational model of neurodegeneration in Alzheimer’s disease. Nat Commun. 2022;13(1):1643.

32. Ramanan S, Halai AD, Garcia-Penton L, et al. The neural substrates of transdiagnostic cognitive-linguistic heterogeneity in primary progressive aphasia. Alzheimers Res Ther. 2023;15(1):219.

33. Johnson JC, McWhirter L, Hardy CJ, et al. Suspecting dementia: canaries, chameleons and zebras. Practical Neurology. 2021;21(4):300–312.

34. Ramanan S, Bertoux M, Flanagan E, et al. Longitudinal Executive Function and Episodic Memory Profiles in Behavioral-Variant Frontotemporal Dementia and Alzheimer’s Disease. J Int Neuropsychol Soc. 2017;23(1):34–43.

35. Graham A, Davies R, Xuereb J, et al. Pathologically proven frontotemporal dementia presenting with severe amnesia. Brain. 2005;128(Pt 3):597–605.

36. Peterson KA, Patterson K, Rowe JB. Language impairment in progressive supranuclear palsy and corticobasal syndrome. Journal of neurology. 2021;268(3):796–809.

37. Cornblath EJ, Robinson JL, Irwin DJ, et al. Defining and predicting transdiagnostic categories of neurodegenerative disease. Nat Biomed Eng. 2020;4(8):787–800.

38. Pini L, de Lange SC, Pizzini FB, et al. A low-dimensional cognitive-network space in Alzheimer’s disease and frontotemporal dementia. Alzheimer’s Research & Therapy. 2022;14(1):199.

39. Beekly DL, Ramos EM, Lee WW, et al. The National Alzheimer’s Coordinating Center (NACC) database: the uniform data set. Alzheimer Disease & Associated Disorders. 2007;21(3):249–258.

40. Morrow CB, Leoutsakos J-MS, Onyike CU. Functional Disabilities and Psychiatric Symptoms in Primary Progressive Aphasia. The American Journal of Geriatric Psychiatry. 2022;30(3):372–382.

41. Mesulam MM. Primary progressive aphasia. Ann Neurol. 2001;49(4):425–432.

42. Cummings JL. The Neuropsychiatric Inventory: assessing psychopathology in dementia patients. Neurology. 1997;48(5 Suppl 6):10S–16S.

43. Yesavage JA. Geriatric depression scale. Psychopharmacol bull. 1988;24(4):709–711.

44. Pfeffer RI, Kurosaki TT, Harrah Jr C, Chance JM, Filos S. Measurement of functional activities in older adults in the community. Journal of gerontology. 1982;37(3):323–329.

45. Morris JC. The Clinical Dementia Rating (CDR): current version and scoring rules. Neurology. 1993;43(11):2412–2414.

46. Knopman DS, Kramer JH, Boeve BF, et al. Development of methodology for conducting clinical trials in frontotemporal lobar degeneration. Brain. 2008;131(Pt 11):2957–2968.

47. Besser LM, Kukull WA, Teylan MA, et al. The Revised National Alzheimer’s Coordinating Center’s Neuropathology Form—Available Data and New Analyses. Journal of Neuropathology & Experimental Neurology. 2018;77(8):717–726.

48. Montine TJ, Phelps CH, Beach TG, et al. National Institute on Aging–Alzheimer’s Association guidelines for the neuropathologic assessment of Alzheimer’s disease: a practical approach. Acta neuropathologica. 2012;123:1–11.

49. R: A language and environment for statistical computing [computer program]. R Foundation for Statistical Computing, Vienna, Austria; 2022.

50. MATLAB [computer program]. Version 7.10.0. Natick, MA. 2010.

51. Qiu Y, Jacobs DM, Messer K, Salmon DP, Feldman HH. Cognitive heterogeneity in probable Alzheimer disease: Clinical and neuropathologic features. Neurology. 2019;93(8):e778–e790.

52. Ballabio D. A MATLAB toolbox for Principal Component Analysis and unsupervised exploration of data structure. Chemometrics and Intelligent Laboratory Systems. 2015;149:1–9.

53. McInnes L, Healy J, Melville J. Umap: Uniform manifold approximation and projection for dimension reduction. arXiv preprint arXiv:180203426. 2018.

54. Han H, Li W, Wang J, Qin G, Qin X. Enhance explainability of manifold learning. Neurocomputing. 2022;500:877–895.

55. Wang Y, Huang H, Rudin C, Shaposhnik Y. Understanding how dimension reduction tools work: an empirical approach to deciphering t-SNE, UMAP, TriMAP, and PaCMAP for data visualization. The Journal of Machine Learning Research. 2021;22(1):9129–9201.

56. Rainer RJ, Mayr M, Himmelbauer J, Nikzad-Langerodi R. Opening the black-box of Neighbor Embeddings with Hotelling’s T2 statistic and Q-residuals. Chemometrics and Intelligent Laboratory Systems. 2023;238:104840.

57. Yang Y, Sun H, Zhang Y, et al. Dimensionality reduction by UMAP reinforces sample heterogeneity analysis in bulk transcriptomic data. Cell reports. 2021;36(4):109442.

58. Diaz-Papkovich A, Anderson-Trocmé L, Gravel S. A review of UMAP in population genetics. Journal of Human Genetics. 2021;66(1):85–91.

59. Langdon C, Genkin M, Engel TA. A unifying perspective on neural manifolds and circuits for cognition. Nature Reviews Neuroscience. 2023;24(6):363–377.

60. Grollemund V, Chat GL, Secchi-Buhour M-S, et al. Development and validation of a 1-year survival prognosis estimation model for Amyotrophic Lateral Sclerosis using manifold learning algorithm UMAP. Scientific reports. 2020;10(1):13378.

61. Kobak D, Linderman GC. Initialization is critical for preserving global data structure in both t-SNE and UMAP. Nature Biotechnology. 2021;39(2):156–157.

62. Kobak D, Berens P. The art of using t-SNE for single-cell transcriptomics. Nature Communications. 2019;10(1):5416.

63. Dadu A, Satone VK, Kaur R, et al. Application of Aligned-UMAP to longitudinal biomedical studies. Patterns. 2022.

64. Leroy M, Bertoux M, Skrobala E, et al. Characteristics and progression of patients with frontotemporal dementia in a regional memory clinic network. Alzheimer’s Research & Therapy. 2021;13(1):19.

65. Giebel CM, Knopman D, Mioshi E, Khondoker M. Distinguishing Frontotemporal Dementia From Alzheimer Disease Through Everyday Function Profiles: Trajectories of Change. J Geriatr Psychiatry Neurol. 2021;34(1):66–75.

66. Devi G, Scheltens P. Heterogeneity of Alzheimer’s disease: consequence for drug trials? Alzheimer’s Research & Therapy. 2018;10(1):1–3.

67. Halai AD, Perez BDD, Stefaniak JD, Ralph MAL. Efficient and effective assessment of deficits and their neural bases in stroke aphasia. cortex. 2022;155:333–346.

68. Ramanan S, de Souza LC, Moreau N, et al. Determinants of theory of mind performance in Alzheimer’s disease: a data-mining study. Cortex. 2017;88:8–18.

69. Green C, Zhang S. Predicting the progression of Alzheimer’s disease dementia: a multidomain health policy model. Alzheimer’s & Dementia. 2016;12(7):776–785.

70. de la Sablonnière J, Tastevin M, Lavoie M, Laforce Jr R. Longitudinal changes in cognition, behaviours, and functional abilities in the three main variants of primary progressive aphasia: A literature review. Brain Sciences. 2021;11(9):1209.

71. Foxe D, Irish M, Ramanan S, et al. Longitudinal changes in behaviour, mood and functional capacity in the primary progressive aphasia variants. Eur J Neurosci. 2021.

72. Pillai JA, Bena J, Maly EF, Leverenz JB. Initial non-amnestic symptoms relate to faster rate of functional and cognitive decline compared to amnestic symptoms in neuropathologically confirmed dementias. Alzheimers Dement. 2023;19(7):2956–2965.

73. Owens TE, Machulda MM, Duffy JR, et al. Patterns of Neuropsychological Dysfunction and Cortical Volume Changes in Logopenic Aphasia. Journal of Alzheimers Disease. 2018;66(3):1015–1025.

74. Hardy CJ, Taylor-Rubin C, Taylor B, et al. Symptom-led staging for semantic and non-fluent/agrammatic variants of primary progressive aphasia. Alzheimer’s & Dementia. 2024;20(1):195–210.

75. Hardy CJ, Taylor-Rubin C, Taylor B, et al. Symptom-based staging for logopenic variant primary progressive aphasia. Eur J Neurol. 2024:e16304.

76. Ulugut H, Stek S, Wagemans LE, et al. The natural history of primary progressive aphasia: beyond aphasia. Journal of neurology. 2022;269(3):1375–1385.

77. Grossman M. Primary progressive aphasia: clinicopathological correlations. Nature Reviews Neurology. 2010;6(2):88–97.

78. Murley AG, Rouse MA, Coyle-Gilchrist IT, et al. Predicting loss of independence and mortality in frontotemporal lobar degeneration syndromes. Journal of Neurology, Neurosurgery & Psychiatry. 2021;92(7):737–744.

79. Libon DJ, Xie SX, Wang X, et al. Neuropsychological decline in frontotemporal lobar degeneration: a longitudinal analysis. Neuropsychology. 2009;23(3):337–346.

80. Lansdall CJ, Coyle-Gilchrist ITS, Jones PS, et al. Apathy and impulsivity in frontotemporal lobar degeneration syndromes. Brain. 2017;140(6):1792–1807.

81. Ducharme S, Price BH, Dickerson BC. Apathy: a neurocircuitry model based on frontotemporal dementia. J Neurol Neurosurg Psychiatry. 2018;89(4):389–396.

82. Ibanez A, Manes F. Contextual social cognition and the behavioral variant of frontotemporal dementia. Neurology. 2012;78(17):1354–1362.

83. Ranasinghe KG, Rankin KP, Lobach IV, et al. Cognition and neuropsychiatry in behavioral variant frontotemporal dementia by disease stage. Neurology. 2016;86(7):600–610.

84. Scarioni M, Gami-Patel P, Peeters CFW, et al. Psychiatric symptoms of frontotemporal dementia and subcortical (co-)pathology burden: new insights. Brain. 2022;146(1):307–320.

85. Kocagoncu E, Klimovich-Gray A, Hughes LE, Rowe JB. Evidence and implications of abnormal predictive coding in dementia. Brain. 2021;144(11):3311–3321.

86. Lansdall CJ, Coyle-Gilchrist ITS, Vázquez Rodríguez P, et al. Prognostic importance of apathy in syndromes associated with frontotemporal lobar degeneration. Neurology. 2019;92(14):e1547–e1557.

87. Rohrer JD, Lashley T, Schott JM, et al. Clinical and neuroanatomical signatures of tissue pathology in frontotemporal lobar degeneration. Brain. 2011;134(9):2565–2581.

88. Warren JD, Rohrer JD, Schott JM, Fox NC, Hardy J, Rossor MN. Molecular nexopathies: a new paradigm of neurodegenerative disease. Trends Neurosci. 2013;36(10):561–569.

89. Warren JD, Fletcher PD, Golden HL. The paradox of syndromic diversity in Alzheimer disease. Nature Reviews Neurology. 2012;8(8):451–464.

90. Hodges JR, Mitchell J, Dawson K, et al. Semantic dementia: demography, familial factors and survival in a consecutive series of 100 cases. Brain. 2010;133(1):300–306.

91. Davenport F, Gallacher J, Kourtzi Z, et al. Neurodegenerative disease of the brain: a survey of interdisciplinary approaches. Journal of the Royal Society Interface. 2023;20(198):20220406.

