## Supplementary for "Mapping the multidimensional geometric landscape of graded phenotypic variation and progression in neurodegenerative syndromes"

**Supplementary Methods**

**Tools and packages used**

The following R packages were used in this project:

Data reading and writing: *data.table*,^1^ *readxl,^2^ openxlsx^3^*

Data cleaning: *tidyverse*,^4^ *reshape2*,^5^ *purrr*,^6^ *stringr*,^7^ *forcats^8^*

Missing data handling: *naniar^9^* (missing data visualisation), *mice*^10^ and *miceadds* (missing data imputation).^10^

Data analysis: *psych* (PCA and post-hoc analyses),^11^ *stats* (Tukey post-hoc analyses and Euclidean distances),^12^ *effectsize* (eta squared effect sizes),^13^ *umap* (UMAP),^14^ *MASS* (2D kernel density estimation),^15^ *lme4* (linear mixed modelling),^16^ *mgcv* (generalised additive models),^17,18^ *ks^19^* (2D kernel density estimation statistics), *sf,^20^ maptools^21^* and *spatstat* (spatial point pattern analysis, average nearest neighbours).^22^

Data visualisation: *ggplot2*,^23^ *ggalluvial*,^24^ *ggrepel*,^25^ *gghighlight*,^26^ *vegan*,^27^ *viridis*,^28^ *cowplot*^29^ (all for static visualisation of data), *gifski^30^* and *gganimate^31^* (animated visualisation of data).

**Characteristics of the PPA group**

The disproportionately greater number of mixed PPA patients, as opposed to canonical variants, appears to be a characteristic of the NACC dataset and has been reported by other groups as well.^32^ We further detailed the characteristics and composition of our PPA group here. Looking at the distribution of our PPA cohort based on the original Alzheimer’s Disease Research Center (ADRC) assigned to their case, of the 17 ADRCs included, the largest number of PPA patients came from centres 289 and 4347 (Supplementary Table 1). We note that the ADRC site numbers are deidentified and, therefore, we cannot ascertain the corresponding geographic location of each site in the USA. The mixed PPA group had a fairly even geographical distribution in terms of centres of origin. These results indicate that our mixed PPA group was not systematically diagnosed in specific centres, rather, emerged fairly evenly across the 17 ADRCs.

**Supplementary Table 1.** Breakdown of our PPA cohort by the Alzheimer’s Disease Research Center number assigned to their case.

| ADRC site number | PPA diagnosis | Count |
| --- | --- | --- |
| 289 | Mixed PPA | 15 |
| 289 | PPA-NOS | 2 |
| 289 | lvPPA | 3 |
| 289 | nfvPPA | 2 |
| 490 | Mixed PPA | 1 |
| 2096 | Mixed PPA | 3 |
| 2096 | svPPA | 1 |
| 2125 | lvPPA | 1 |
| 2289 | Mixed PPA | 1 |
| 2578 | Mixed PPA | 4 |
| 2958 | Mixed PPA | 2 |
| 2958 | lvPPA | 2 |
| 2958 | nfvPPA | 2 |
| 3697 | Mixed PPA | 1 |
| 4347 | Mixed PPA | 12 |
| 4347 | PPA-NOS | 3 |
| 4347 | lvPPA | 8 |
| 4347 | nfvPPA | 8 |
| 4347 | svPPA | 3 |
| 4967 | Mixed PPA | 2 |
| 4967 | PPA-NOS | 2 |
| 4967 | svPPA | 1 |
| 5452 | Mixed PPA | 3 |
| 5783 | lvPPA | 1 |
| 5783 | nfvPPA | 1 |
| 6518 | Mixed PPA | 1 |
| 8354 | Mixed PPA | 1 |
| 8354 | lvPPA | 3 |
| 8354 | svPPA | 2 |
| 8361 | Mixed PPA | 2 |
| 8361 | PPA-NOS | 1 |
| 8658 | Mixed PPA | 1 |
| 9637 | Mixed PPA | 8 |

*Note*. ADRC, Alzheimer’s Disease Research Center; PPA, Primary Progressive Aphasia; NOS, Not Otherwise Specified; lv, logopenic variant; nfv, nonfluent variant; sv, semantic variant.

Next, to understand whether our PPA patients were systematically diagnosed within a specific time range, we looked at the date of entry of our PPA cohort into our NACC data cut. For this, we used the *VISITYR* variable in the NACC data, which corresponds to the year of visit of the patient as entered on the NACC UDS form. The results are displayed in Supplementary Table 2 and indicate that our mixed PPA group were entered into the NACC database between 2005-2015, while the canonical PPA variants were entered into our data cut only in 2015 and onwards.

**Supplementary Table 2.** Breakdown of number of PPA patients, stratified by group and year, entered into our NACC data cut.

| **Year of entry** | **Mixed PPA** | **PPA-NOS** | **lvPPA** | **nfvPPA** | **svPPA** |
| --- | --- | --- | --- | --- | --- |
| 2005 | 1 |  |  |  |  |
| 2006 | 5 |  |  |  |  |
| 2007 | 7 |  |  |  |  |
| 2008 | 8 |  |  |  |  |
| 2009 | 4 |  |  |  |  |
| 2010 | 8 |  |  |  |  |
| 2011 | 7 |  |  |  |  |
| 2012 | 4 |  |  |  |  |
| 2013 | 2 |  |  |  |  |
| 2014 | 9 |  |  |  |  |
| 2015 | 2 | 1 | 4 | 1 | 1 |
| 2016 |  | 2 | 2 | 2 | 1 |
| 2017 |  | 2 | 3 | 4 | 2 |
| 2018 |  | 2 | 6 | 2 | 1 |
| 2019 |  |  | 3 | 4 | 2 |

*Note*. PPA, Primary Progressive Aphasia; NOS, Not Otherwise Specified; lv, logopenic variant; nfv, nonfluent variant; sv, semantic variant.

Next, we looked at the above distribution, but stratified before and after the year 2011, corresponding to the publication of the Gorno-Tempini et al. PPA criteria that were employed in this study for PPA diagnosis. These results are displayed in Supplementary Table 3. These results indicate that all the canonical variants and PPA-NOS patients included in our analyses visited the ADRCs and were entered into the NACC database only post-2011 (corresponding to the Gorno-Tempini et al. PPA criteria publication). In contrast, the majority of our mixed PPA group (40/57, 70%) were entered into the NACC database before 2011.

**Supplementary Table 3.** Breakdown of number of PPA patients entered into our NACC data cut, stratified by pre- and post-2011.

| PPA diagnosis | Entered into the NACC data cut before/after 2011 | Count |
| --- | --- | --- |
| Mixed PPA | Pre-2011 | 40 |
| Mixed PPA | Post-2011 | 17 |
| PPA-NOS | Post-2011 | 8 |
| lvPPA | Post-2011 | 18 |
| nfvPPA | Post-2011 | 13 |
| svPPA | Post-2011 | 7 |

*Note*. PPA, Primary Progressive Aphasia; NOS, Not Otherwise Specified; lv, logopenic variant; nfv, nonfluent variant; sv, semantic variant.

We then looked deeper into the mixed PPA cohort and explored their corresponding diagnosis per the Mesulam et al. (2001, 2003) criteria. To recap, while our PPA cohort was diagnosed as per the current Gorno-Tempini et al. (2011) criteria (corresponding to variable *NACCPPAG*), the NACC data variables also had a variable titled *NACCPPME* corresponding to “PPA subtype according to older criteria outlined by Mesulam et al. (2001 and 2003)”. In the NACC UDS data dictionary, the possible scores for the *NACCPPME* variable include:

- Score of 1 = Meets criteria for progressive nonfluent PPA
- Score of 2 = Meets criteria for semantic dementia – anomia plus word comprehension
- Score of 3 = Meets criteria for semantic dementia – agnostic variant
- Score of 4 = Meets criteria for PPA-Other/Not Otherwise Specified (logopenic, anomic, transcortical, word deafness, syntactic comprehension, motor speech disorder)
- Score of 6 = Subject had MCI but missing information on presence/absence of PPA
- Score of 7 = Subject was cognitively impaired but did not have PPA
- Score of 8 = No cognitive impairment
- Score of -4 = Not applicable

We cross-referenced available *NACCPPME* classification data for our PPA cohort with their Gorno-Tempini diagnostic label. The results indicated that the Mesulam et al. classifications of scores 1-4 were only applied for our mixed PPA cohort. For all patients labelled as semantic, logopenic, nonfluent PPA, and PPA-NOS as per the Gorno-Tempini criteria, the corresponding values in the *NACCPPME* variable were -4 (i.e., Not applicable). These individuals only had scores in the *NACCPPAG* (Gorno-Tempini criteria) column. As the Mesulam et al. classification had only been applied to the mixed PPA patients, we broke down this data for visualisation using a pie chart (Supplementary Figure 1). The results suggest that the majority of individuals classified as mixed PPA in our cohort met the classification for Progressive Nonfluent PPA as per the Mesulam et al. description.^33^ As per the Mesulam et al. (2001, 2003) labels applied in our NACC data cut, the second and third largest composition of our mixed PPA cohort were semantic dementia (anomia and word comprehension) and semantic dementia – agnostic variant.

**Supplementary Figure 1.** Percentage breakdown of our mixed PPA cohort as per application of the Mesulam et al. PPA classification.

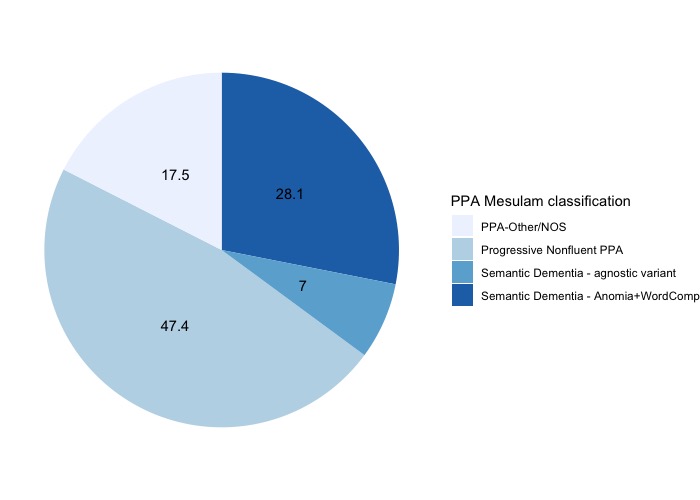

*Note*. PPA, Primary Progressive Aphasia.

**Supplementary Results**

***Global dispersion index changes in individuals with stable vs. changing diagnoses***

Probing the direction of movement within the multidimensional PC space, we split the data by individuals with stable vs. changing diagnostic labels and ran our standard linear mixed-effect model pipeline (fixed effect for group and follow-up time, random effect for follow-up time, a random intercept for individual performance and a random slope for follow-up time). For analysis in the dataset of individuals with changing diagnostic labels, we added an additional random effect of group. In individuals with stable diagnoses, linear mixed-effect models revealed the largest time*group interaction effect in PPA (*t*=-2; *p*=.043). In individuals who changed diagnostic labels, only main effects for time (*t*=-2.2; *p*=.02) and group (AD: *t*=2.5; *p*=.01; PPA: *t*=5.4; p<.001) emerged.

**Supplementary Table 4.** Coded vascular change categories and respective NACC variables.

| **Category of vascular change** | **NACC variable name** |
| --- | --- |
| *Amyloid angiopathy* |  |
| Cerebral amyloid angiopathy | NACCAMY |
| *Non-amyloid angiopathy* |  |
| Non-amyloid angiopathy | NPOANG |
| *Vascular changes* |  |
| Large arterial infarcts | NPLINF; NACCINF |
| One/more lacunes | NPLAC; NACCINF |
| Microinfarcts | NPMICRO; NPOLD; NACCMICR |
| Arteriosclerosis | NACCARTE |
| Subcortical arteriosclerotic leukoencephalopathy | NPART |
| White matter rarefaction | NPWMR |
| Other pathologic changes related to ischemic or vascular disease not previously specified | NPPATH |
| Laminar necrosis | NACCNEC |
| Mineralization of blood vessels | NPPATH11 |
| Other ischemic/vascular pathology | NPPATH0 |
| *Acute injury* |  |
| Single/multiple haemorrhages | NPHEM; NPHEMO; NACCHEM |
| Cerebral microbleeds | NPOLDD; NACCHEM |
| Acute neuronal necrosis | NPPATH2 |
| Acute/subacute gross infarcts | NPPATH3 |
| Acute/subacute microinfarcts | NPPATH4 |
| Acute/subacute gross haemorrhage | NPPATH5 |
| Acute/subacute microhaemorrhage | NPPATH6 |

Note. NACC=National Alzheimer’s Coordinating Centre dataset.

**Supplementary Table 5**. Two tailed Pearson’s correlation *r* values between global dispersion index and disease severity as measured by the CDR-FTLD-SoB in all patient groups (irrespective of visit).

| **Group** | **Correlation statistic** |
| --- | --- |
| AD | *r*=-.58; *p*<.001 |
| bvFTD | *r*=-.22; *p*<.001 |
| FTLD-Motor | *r*=-.33; *p*<.001 |
| FTLD-NoS | *r*=-.42; *p*<.001 |
| PPA | *r*=-.59; *p*<.001 |

*Note*. AD=Alzheimer’s disease; bvFTD=behavioural variant frontotemporal dementia; FTLD=frontotemporal lobar degeneration; NOS=not otherwise specified; PPA=primary progressive aphasia; CDR-FTLD-SoB=Clinical Dementia Rating Plus NACC FTLD Sum of Boxes.

**Supplementary Table 6**. Statistical differences between spatial locations occupied by different groups at each time point in the UMAP space.

|  | **AD** | **bvFTD** | **FTLD-Motor** | **FTD-NOS** | **PPA** |
| --- | --- | --- | --- | --- | --- |
| Visit 1 vs. 2 | *Z*=.3; *p*=.3 | *Z*=.7; *p*=.2 | *Z*=-.1; *p*=.5 | *Z*=1.2; *p*=.1 | ***Z*=7.2; *p*<.001** |
| Visit 2 vs. 3 | *Z*=.3; *p*=.3 | *Z*=-.4; *p*=.6 | *Z*=.1; *p*=.4 | *Z*=-.2; *p*=.5 | *Z*=1.3; *p*=.08 |
| Visit 1 vs. 3 | ***Z*=2.4; *p*=.008** | ***Z*=3.2; *p*<.001** | ***Z*=1.8; *p*=.03** | ***Z*=2.3; *p*=.008** | ***Z*=13.3; *p*<.001** |

*Note*. Statistical differences tested using spatial kernel density estimation statistics. AD=Alzheimer’s disease; bvFTD=behavioural variant frontotemporal dementia; FTLD=frontotemporal lobar degeneration; NOS=not otherwise specified; PPA=primary progressive aphasia.

**Supplementary Table 7.** Average nearest neighbour analysis for observed versus null distributions for each patient group in the UMAP space.

|  | Visit 1 | Visit 2 | Visit 3 |
| --- | --- | --- | --- |
| AD | **ANN=.26; *p*=.029** | ANN=.28; *p*=.10 | ANN=.25; *p*=.11 |
| bvFTD | **ANN =.26; *p*=.009** | **ANN=.31; *p*=.009** | ANN=.34; *p*=.43 |
| FTLD-motor | **ANN=.37; *p*=.001** | **ANN=.51; *p*=.026** | **ANN=.39; *p*=.002** |
| FTLD-NOS | ANN=.6; *p*=.19 | **ANN=.52; *p*=.009** | ANN=.58; *p*=.2 |
| PPA | ANN=.32; *p*=.31 | ANN=.31; *p*=.08 | ANN=.29; *p*=.14 |

*Note.* ANN=average nearest neighbour value; *p*-values derived from statistical comparison of observed data to a null distribution that is derived by randomly shuffling diagnostic labels. AD=Alzheimer’s disease; bvFTD=behavioural variant frontotemporal dementia; FTLD=frontotemporal lobar degeneration; NOS=not otherwise specified; PPA=primary progressive aphasia.

**Supplementary Table 8.** Relationship between PC-specific centroids and survival at 36 months

|  | AD | FTLD-Motor | PPA |
| --- | --- | --- | --- |
| *PC1 (Functional status)* |  |  |  |
| Starting point | *Z*=-1.9; *p*=.04 | *Z*=-2.3; *p*=.01 |  |
| Travelled distance |  |  | *Z*=-2.4; *p*=.01 |
| *PC3 (Motor function)* |  |  |  |
| Starting point |  | *Z*=2.5; *p*=.01 |  |
| Travelled distance |  | *Z*=-2.2; *p*=.02 | *Z*=-2.2; *p*=.02 |
| *PC4 (Psychosis)* |  |  |  |
| Starting point | *Z*=-2; *p*=.04 |  |  |
| Travelled distance |  |  |  |
| *PC5 (Affective changes)* |  |  |  |
| Starting point |  |  | *Z*=2.6; *p*=.008 |
| Travelled distance |  |  |  |

*Note*. No significant associations emerged for the bvFTD group and the model did not converge for the FTLD-NOS due to small sample size.

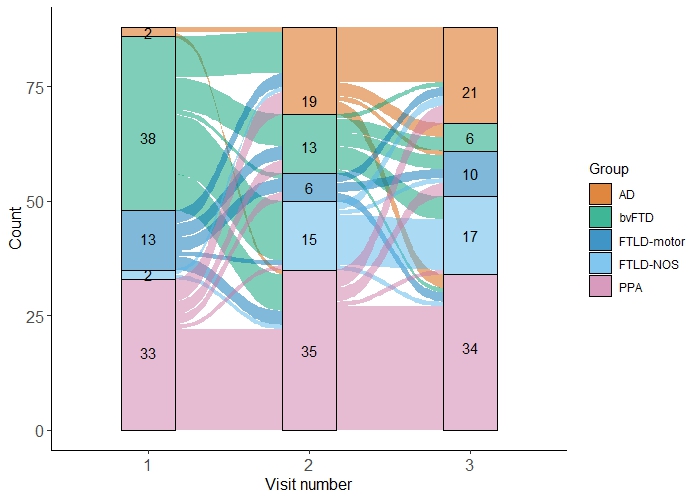

**Supplementary Figure 2.** Alluvial plot indicating number of individuals changing diagnostic labels during subsequent NACC visits. NACC=National Alzheimer’s Coordinating Centre; AD=Alzheimer’s disease; bvFTD=behavioural variant frontotemporal dementia; FTLD=frontotemporal lobar degeneration; NOS=not otherwise specified; PPA=primary progressive aphasia.

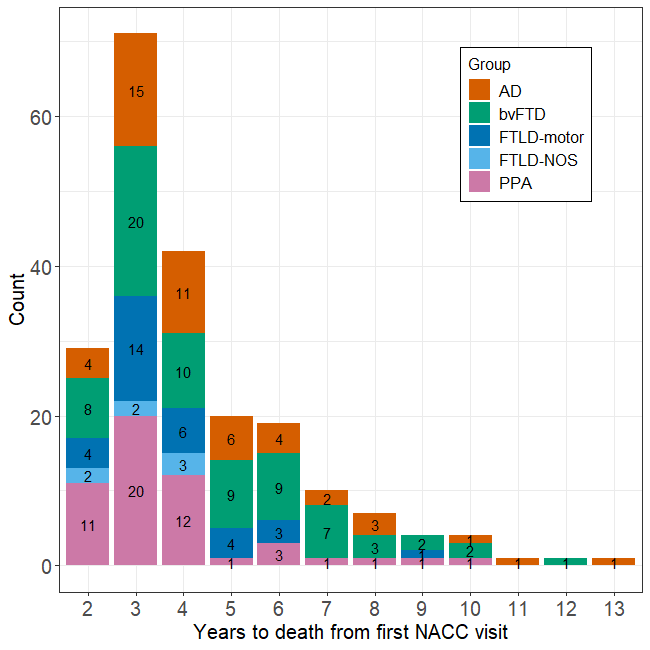

**Supplementary Figure 3. Years to death from first reported visit in our sample from the NACC dataset.** AD=Alzheimer’s disease; bvFTD=behavioural variant frontotemporal dementia; FTLD=frontotemporal lobar degeneration; NOS=not otherwise specified; PPA=primary progressive aphasia; NACC=National Alzheimer’s Coordinating Centre.

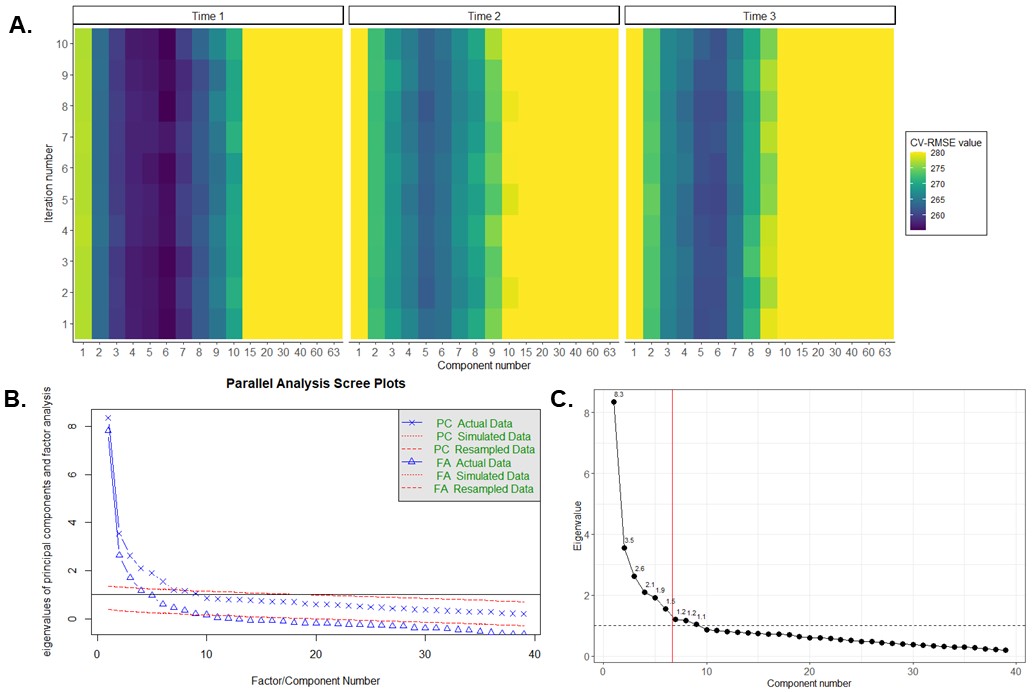

**Supplementary Figure 4.** Determining optimal number of principal components underlying the full dataset (*N*=1,170). Panel **A)** Four-fold cross-validated PCA component selection on the dataset stratified by visit. Colours correspond to cross-validated root-mean-squared-error (CV-RMSE) values for optimal component solution (*x*-axis) across 10 iterations of the algorithm (*y*-axis). The average number of components with the lowest underlying CV-RMSE values across all three time points (here, 6) represents the optimal solution. CV-RMSE values above 280 are truncated for visualisation. Panel **B)** Parallel analysis on the full dataset (not stratified by visit) indicating ideal component solution (here, 6) as displayed by number of X symbols crossing the PC simulated data line (top red dotted line). Panel **C)** Eigenvalue scree plot for the on the full dataset (not stratified by visit) indicating number of components with eigenvalue>1, with vertical red line indicating a 6 component cut-off. PCA=principal component analysis.

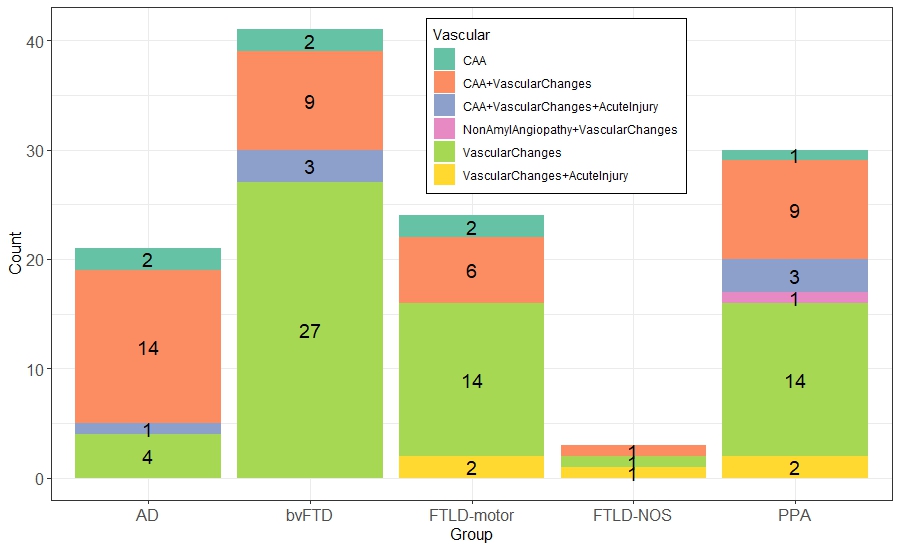

**Supplementary Figure 5.** Frequency of vascular changes in each clinical diagnostic group. AD=Alzheimer’s disease; bvFTD=behavioural variant frontotemporal dementia; FTLD=frontotemporal lobar degeneration; NOS=not otherwise specified; PPA=primary progressive aphasia; CAA=cerebral amyloid angiopathy.

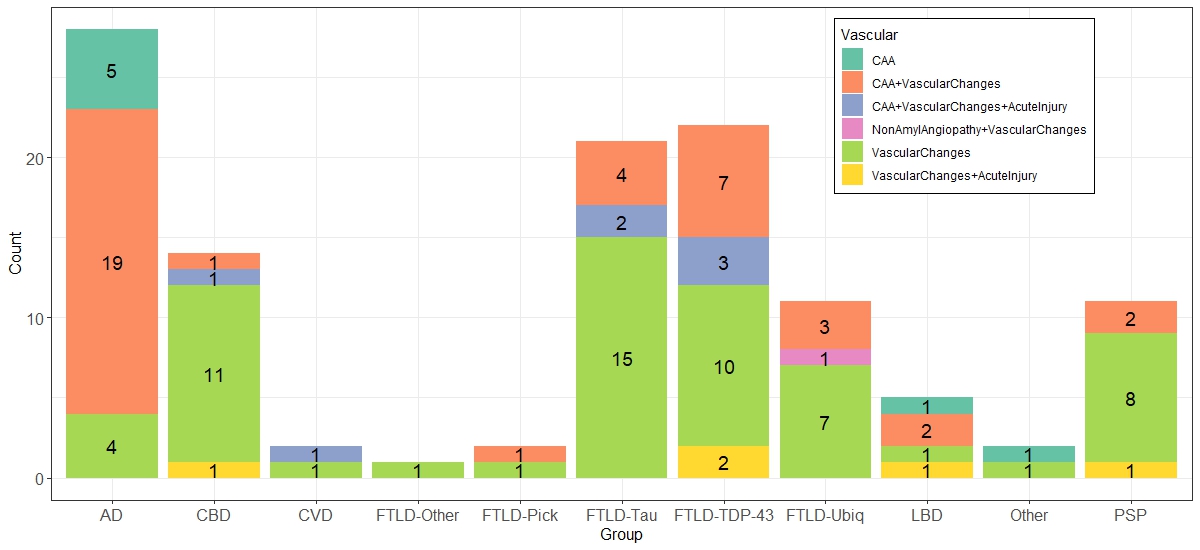

**Supplementary Figure 6.** Frequency of vascular changes in each pathological diagnostic group. AD=Alzheimer’s disease; FTLD=frontotemporal lobar degeneration; TDP-43=Tar DNA binding protein-43; LBD=Lewy Body Disease; CBD=Corticobasal Degeneration; CVD=cardiovascular disease; Ubiq=ubiquitin; PSP=Progressive Supranuclear Palsy; CAA=cerebral amyloid angiopathy.

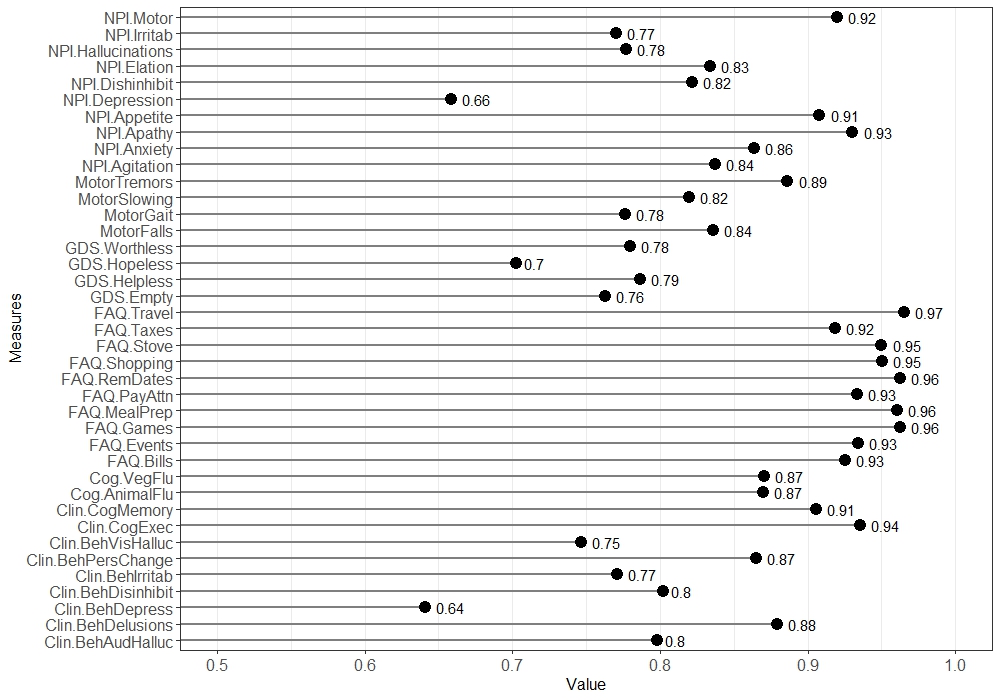

**Supplementary Figure 7.** Measurement of sampling adequacy for individual items included in the principal component analysis. NPI=Neuropsychiatric Inventory; GDS=Geriatric Depression Scale; FAQ=Functional Activities Questionnaire; Clin=Clinical; Beh=Behavioural; Cog=Cognitive; Flu=Fluency.

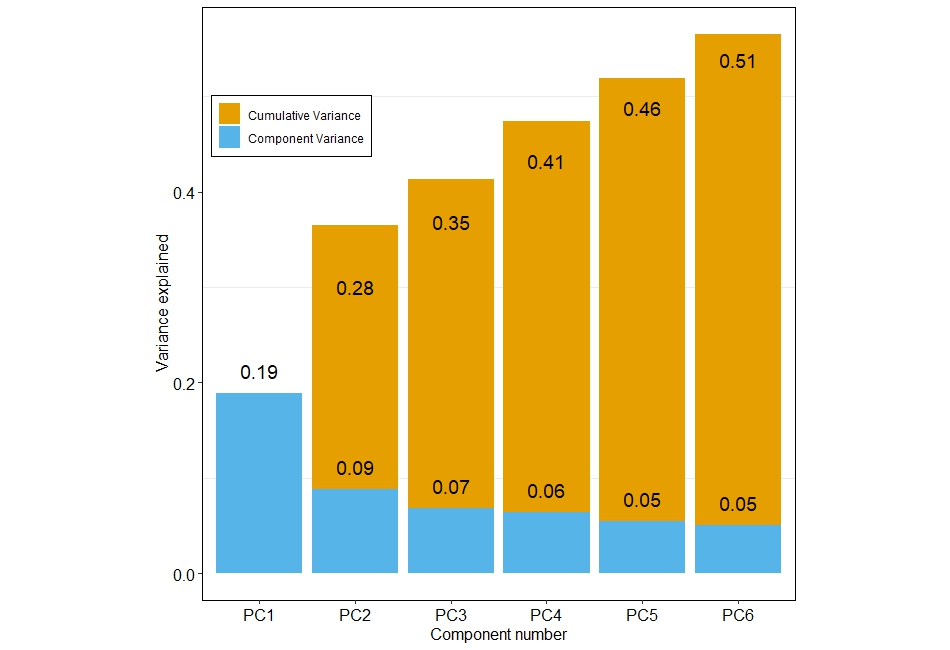

**Supplementary Figure 8.** Cumulative and component-specific variance explained by a six component PCA solution. PCA=principal component analysis.

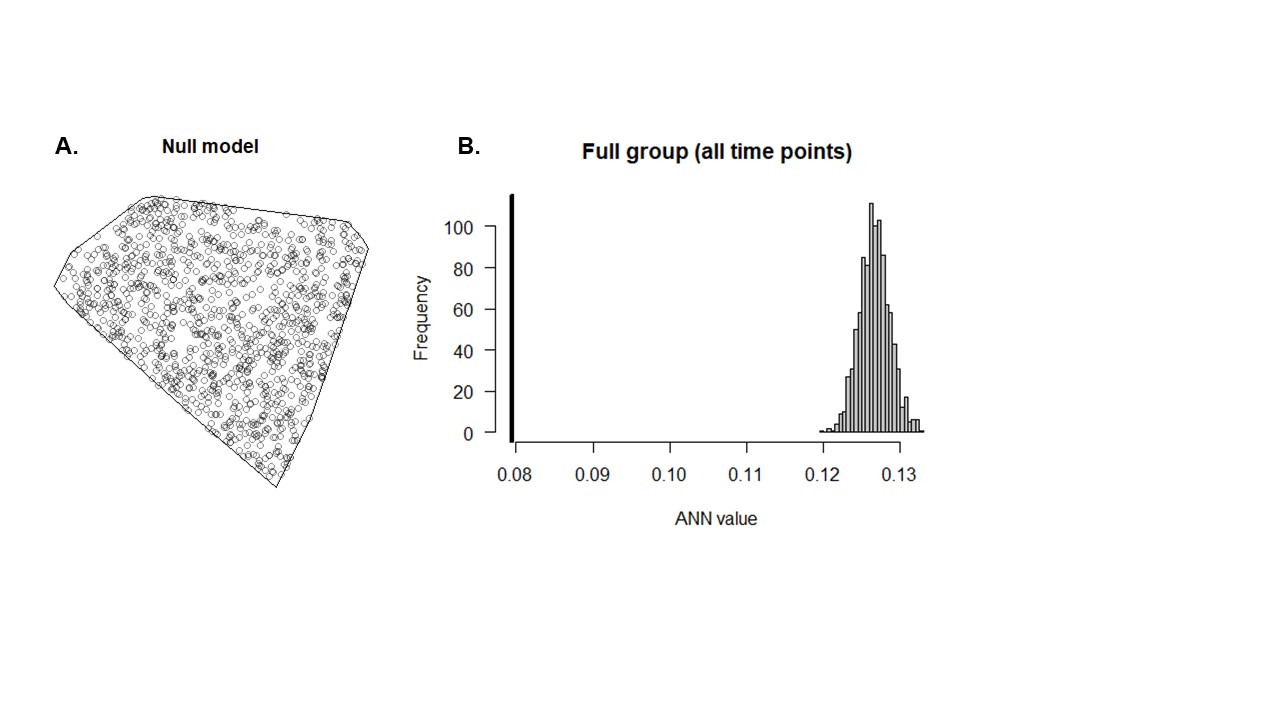

**Supplementary Figure 9. Average Nearest Neighbours analyses in the full group (all time points) to test the statistical structure of the UMAP embedding**. Panel **A)** Null model of 1,170 points created by sampling random points enclosed within the convex hull of our UMAP space. Panel **B)** Histogram of null distribution of average nearest neighbours analysis (mean null value=.12) compared to our observed average nearest neighbour value from the UMAP embedding (black bar; mean observed value=.07). The observed value is much smaller than one expected under the null hypothesis suggesting our UMAP embedding follows a non-random spatial distribution. ANN=average nearest neighbours; UMAP=Uniform Manifold Approximation and Projection.

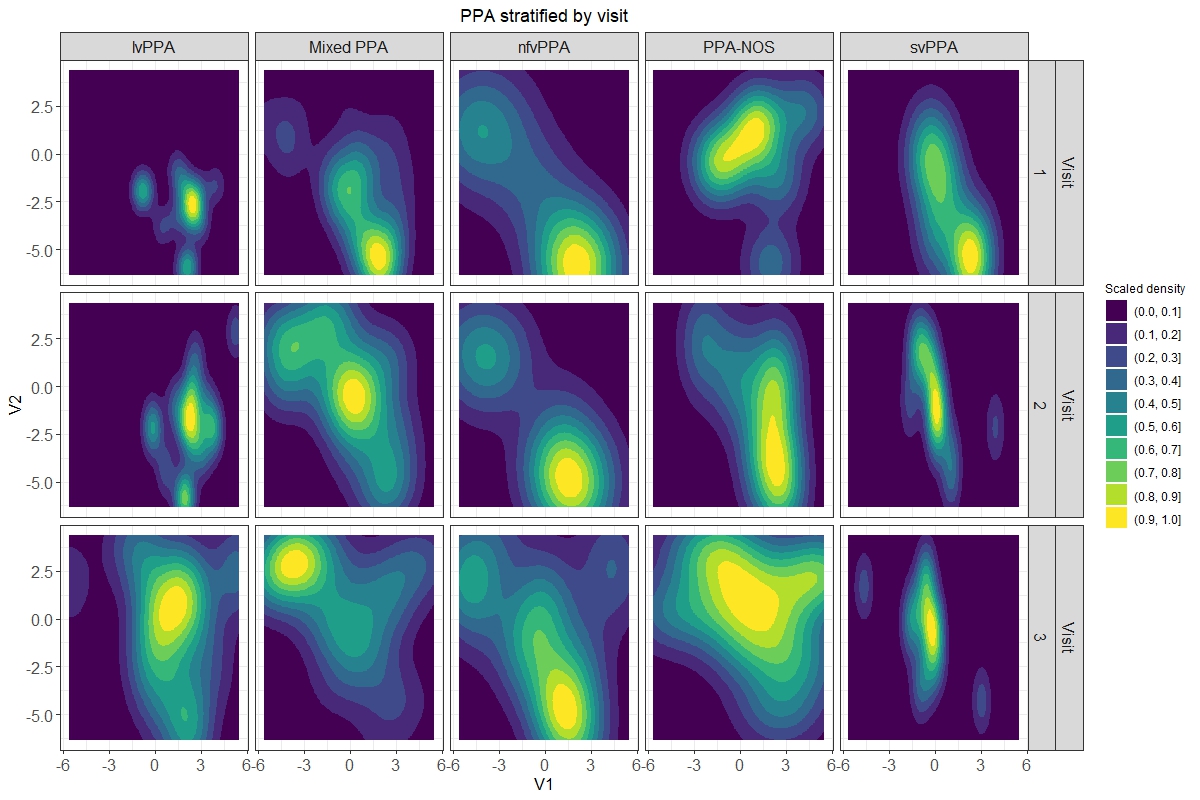

**Supplementary Figure 10. Spatial kernel density estimation plots indicating the “movement” of PPA groups within the UMAP space over time.** Warm colours indicate the greatest sample density, scaled to the overall sample size. PPA=primary progressive aphasia; lv=logopenic variant; nfv=nonfluent variant; NOS=not otherwise specified; sv=semantic variant; UMAP=Uniform Manifold Approximation and Projection.

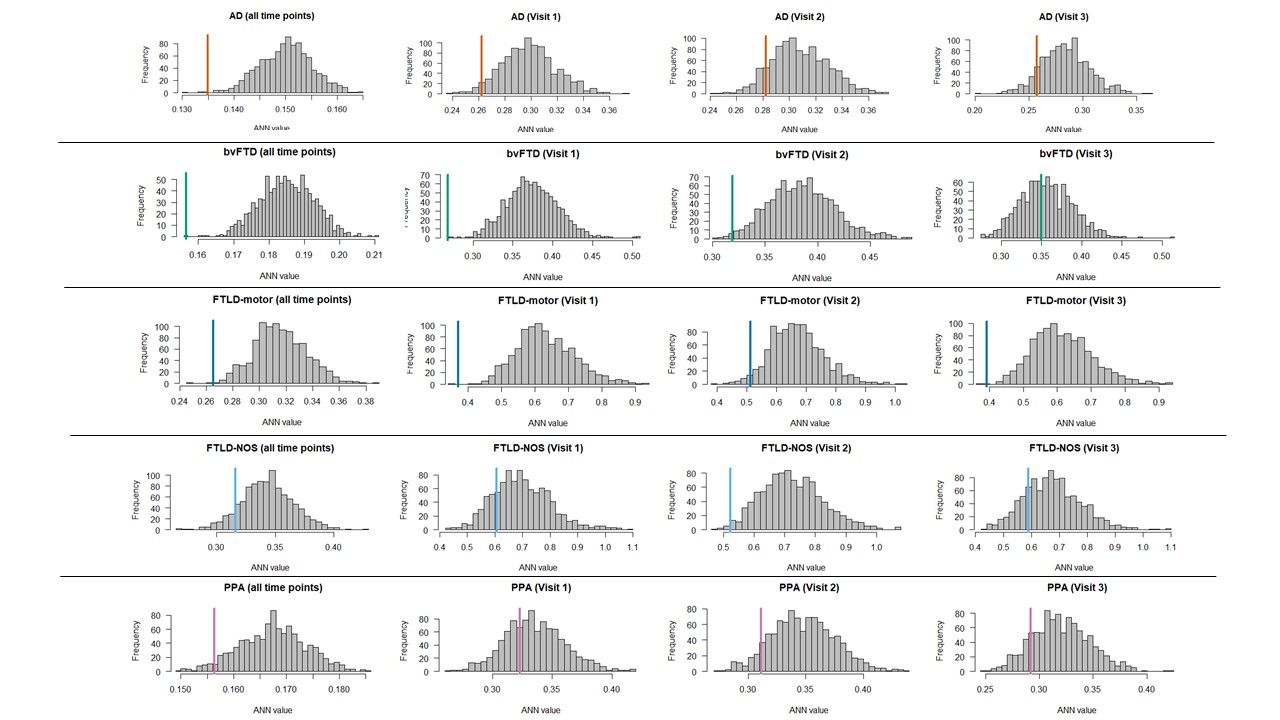

**Supplementary Figure 11. Average Nearest Neighbours analyses in each patient group, stratified by time, to test the statistical structure of group-wise embedding in the UMAP space.** Histograms showcase null models generated for each stratified dataset, created by randomly shuffling diagnostic labels sampled equivalent to the sample size of each stratified dataset. Horizontal bars indicated observed average nearest neighbour values respective to the null distribution. ANN=average nearest neighbours; AD=Alzheimer’s disease; bvFTD=behavioural variant frontotemporal dementia; FTLD=frontotemporal lobar degeneration; NOS=not otherwise specified; PPA=primary progressive aphasia.

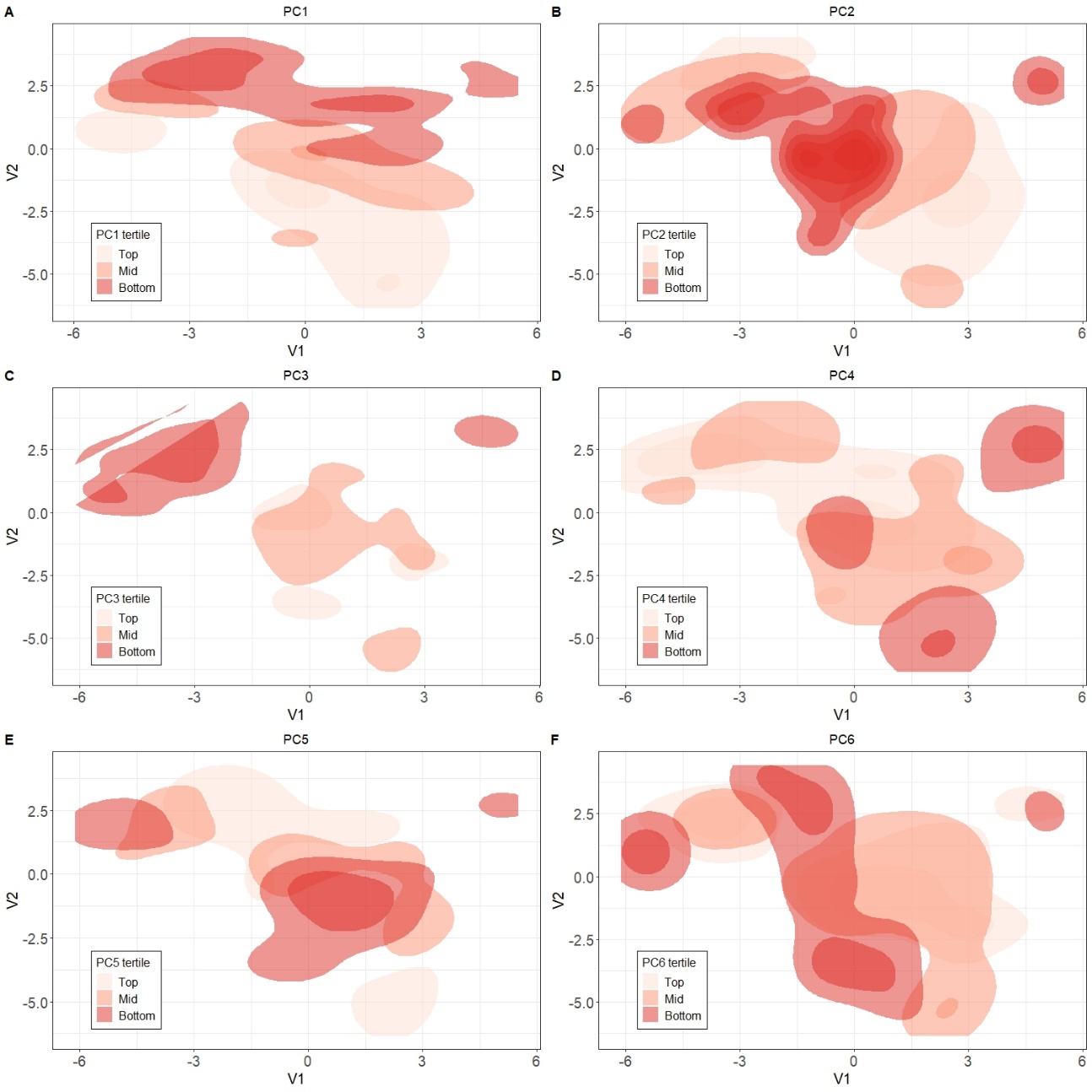

**Supplementary Figure 12. Tertile kernel density plots for all PCs projected into the UMAP space.** Tertiles are represented as 2D kernel density estimate contours. PC=principal component; UMAP=Uniform Manifold Approximation and Projection.

**Supplementary Figure 13. Predominant symptom first recognised as a decline in each patient’s behaviour projected onto UMAP space.** The NACCBEHF assessment can be scored across 10 domains including agitation, apathy/withdrawal, depressed mood, disinhibition, irritability, no behavioural symptoms, other, personality changes, psychosis and REM sleep behavioural disorder. Panel A) displays the data projected into the UMAP space, whereas panel B) displays the accompanying frequency counts of this data (values jittered to prevent overlap). AD=Alzheimer’s disease; bvFTD=behavioural variant frontotemporal dementia; FTLD=frontotemporal lobar degeneration; NOS=not otherwise specified; PPA=primary progressive aphasia; UMAP=Uniform Manifold Approximation and Projection.

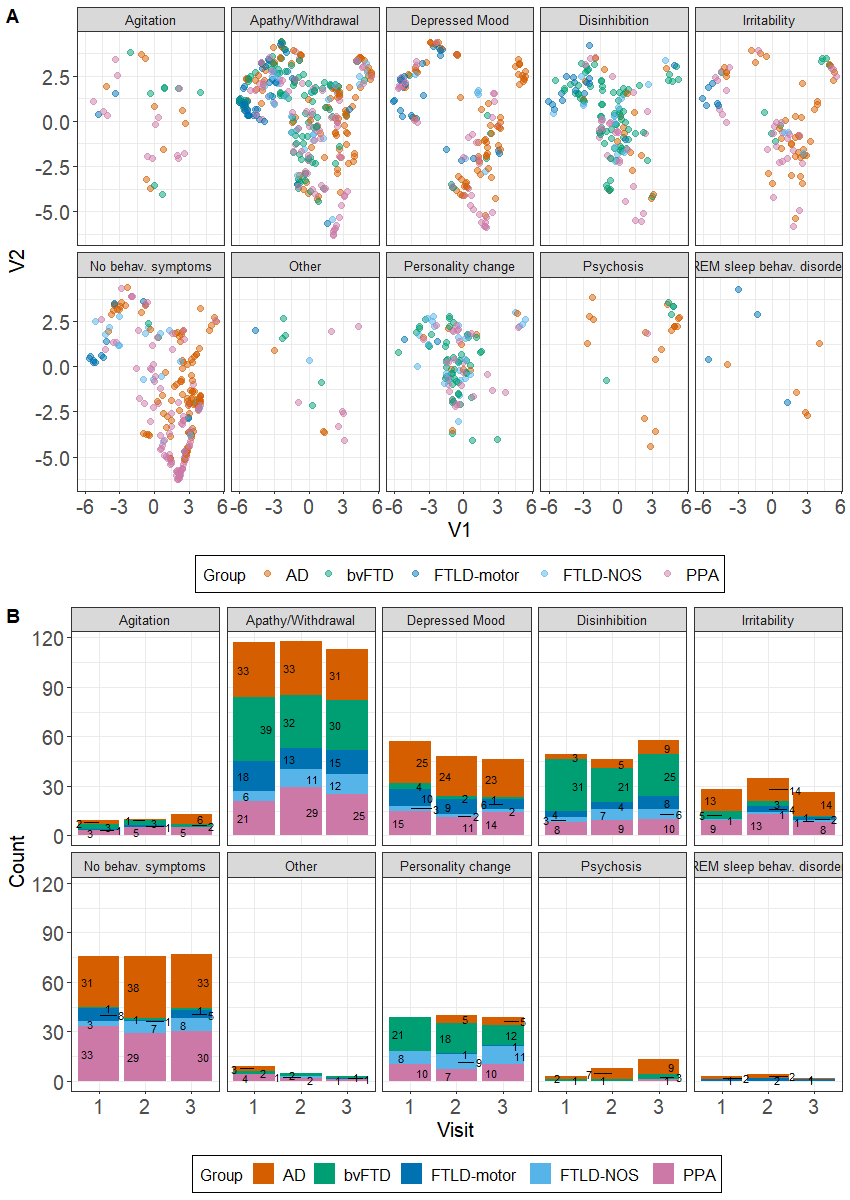

**Supplementary Figure 14. Predominant symptom first recognised as a decline in each patient’s cognition projected onto UMAP space.** The NACCCOGF measure can be scored across eight domains including attention/concentration, executive functions, language, memory, no cognitive symptoms, orientation, other, and visuospatial functions. Panel A) displays the data projected into the UMAP space, whereas panel B) displays the accompanying frequency counts of this data (values jittered to prevent overlap). AD=Alzheimer’s disease; bvFTD=behavioural variant frontotemporal dementia; FTLD=frontotemporal lobar degeneration; NOS=not otherwise specified; PPA=primary progressive aphasia; UMAP=Uniform Manifold Approximation and Projection.

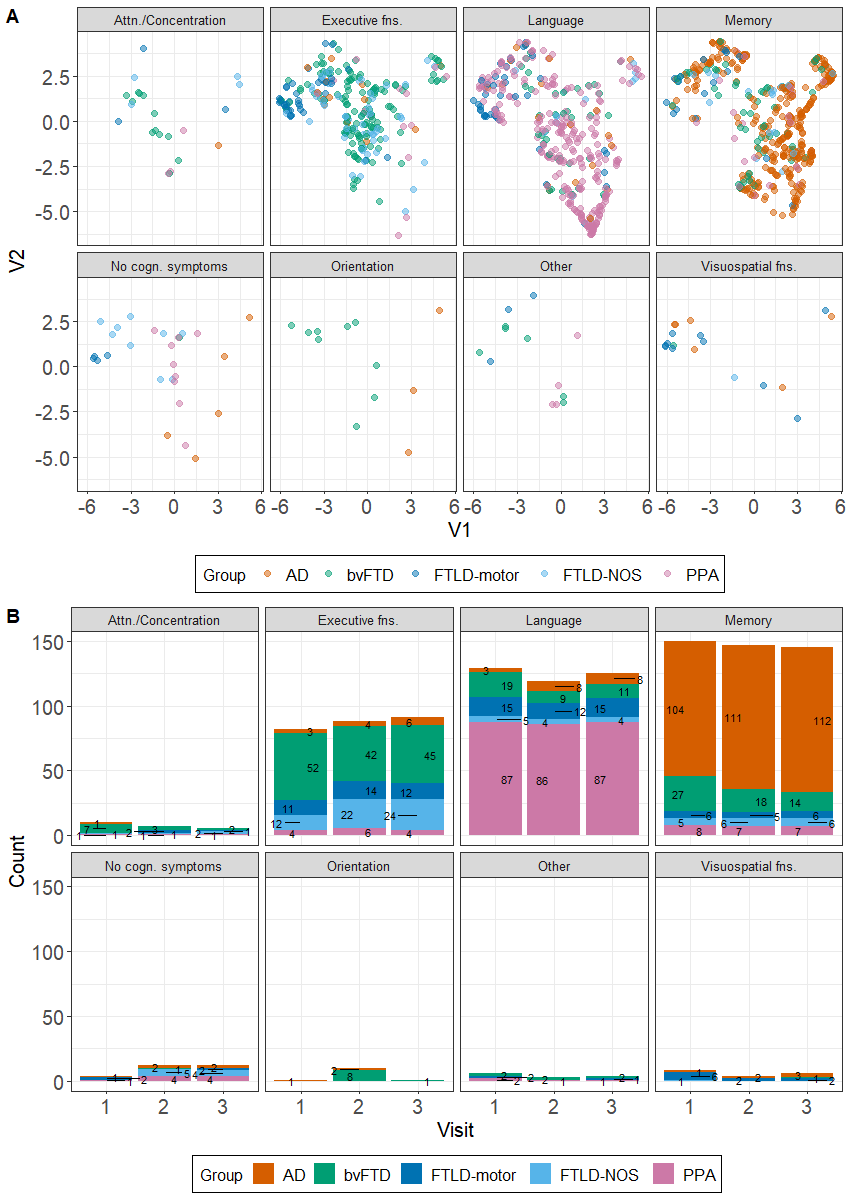

**Supplementary References**

1.	*data.table: Extension of `data.frame`* [computer program]. Version R package version 1.14.22021.

2.	*readxl: Read Excel Files* [computer program]. Version R package version 1.4.22023.

3.	*openxlsx: Read, Write and Edit xlsx Files* [computer program]. Version R package version 4.2.5.12022.

4.	Wickham H, Averick M, Bryan J, et al. Welcome to the Tidyverse. *Journal of open source software.* 2019;4(43):1686.

5.	Wickham H. Reshaping Data with the reshape Package. *Journal of Statistical Software.* 2007;21(12):1 - 20.

6.	*purrr: Functional Programming Tools* [computer program]. Version R package version 1.0.12023.

7.	*stringr: Simple, Consistent Wrappers for Common String Operations* [computer program]. Version R package version 1.5.02022.

8.	*forcats: Tools for Working with Categorical Variables (Factors)* [computer program]. Version R package version 1.0.02023.

9.	Tierney N, Cook D. Expanding Tidy Data Principles to Facilitate Missing Data Exploration, Visualization and Assessment of Imputations. *Journal of Statistical Software.* 2023;105(7):1 - 31.

10.	Van Buuren S, Groothuis-Oudshoorn K. mice: Multivariate imputation by chained equations in R. *Journal of statistical software.* 2011;45:1-67.

11.	Revelle WR. psych: Procedures for personality and psychological research. 2017.

12.	*R: A language and environment for statistical computing* [computer program]. R Foundation for Statistical Computing, Vienna, Austria; 2022.

13.	Ben-Shachar MS, Lüdecke D, Makowski D. effectsize: Estimation of effect size indices and standardized parameters. *Journal of Open Source Software.* 2020;5(56):2815.

14.	*umap: Uniform Manifold Approximation and Projection* [computer program]. Version R package version 0.2.9.02022.

15.	Venables WN, Ripley BD. *Modern applied statistics with S-PLUS.* Springer Science & Business Media; 2013.

16.	Bates D, Mächler M, Bolker B, Walker S. Fitting linear mixed-effects models using lme4. *arXiv preprint arXiv:14065823.* 2014.

17.	Wood SN. Fast stable restricted maximum likelihood and marginal likelihood estimation of semiparametric generalized linear models. *Journal of the Royal Statistical Society Series B: Statistical Methodology.* 2011;73(1):3-36.

18.	Wood SN. Thin plate regression splines. *Journal of the Royal Statistical Society Series B: Statistical Methodology.* 2003;65(1):95-114.

19.	Duong T. ks: Kernel density estimation and kernel discriminant analysis for multivariate data in R. *Journal of statistical software.* 2007;21:1-16.

20.	Pebesma EJ. Simple features for R: standardized support for spatial vector data. *R J.* 2018;10(1):439.

21.	*maptools: Tools for Handling Spatial Objects* [computer program]. Version R package version 1.1-62022.

22.	Baddeley A, Rubak E, Turner R. *Spatial point patterns: methodology and applications with R.* CRC press; 2015.

23.	*ggplot2: Elegant Graphics for Data Analysis* [computer program]. New York: Springer-Verlag; 2016.

24.	Brunson JC. Ggalluvial: layered grammar for alluvial plots. *Journal of Open Source Software.* 2020;5(49).

25.	*ggrepel: Automatically Position Non-Overlapping Text Labels with 'ggplot2'* [computer program]. Version R package version 0.9.22022.

26.	*gghighlight: Highlight Lines and Points in 'ggplot2'* [computer program]. Version R package version 0.4.02022.

27.	Oksanen J, Kindt R, Legendre P, et al. The vegan package. *Community ecology package.* 2007;10(631-637):719.

28.	*Rvision - Colorblind-Friendly Color Maps for R.* [computer program]. Version R package version 0.6.22021.

29.	*cowplot: Streamlined Plot Theme and Plot Annotations for 'ggplot2'* [computer program]. Version R package version 1.1.12020.

30.	*gifski: Highest Quality GIF Encoder* [computer program]. Version R package version 1.6.6-12022.

31.	*gganimate: A Grammar of Animated Graphics* [computer program]. Version R package version 1.0.82022.

32.	Morrow CB, Leoutsakos J-MS, Onyike CU. Functional Disabilities and Psychiatric Symptoms in Primary Progressive Aphasia. *The American Journal of Geriatric Psychiatry.* 2022;30(3):372-382.

33.	Mesulam MM. Primary progressive aphasia. *Ann Neurol.* 2001;49(4):425-432.
